# Drug-induced epigenomic plasticity reprograms circadian rhythm regulation to drive prostate cancer towards androgen-independence

**DOI:** 10.1101/2021.11.02.21265806

**Authors:** Simon Linder, Marlous Hoogstraat, Suzan Stelloo, Karianne Schuurman, Hilda de Barros, Maartje Alkemade, Joyce Sanders, Yongsoo Kim, Elise Bekers, Jeroen de Jong, Roelof J.C. Kluin, Claudia Giambartolomei, Ji-Heui Seo, Bogdan Pasaniuc, Amina Zoubeidi, Matthew L. Freedman, Lodewyk F.A. Wessels, Lisa M. Butler, Nathan A. Lack, Henk van der Poel, Andries M. Bergman, Wilbert Zwart

## Abstract

In prostate cancer, androgen receptor (AR)-targeting agents are very effective in various stages of the disease. However, therapy resistance inevitably occurs and little is known about how tumor cells adapt to bypass AR suppression. Here, we performed integrative multi-omics analyses on tissues isolated before and after 3 months of AR-targeting enzalutamide monotherapy from high-risk prostate cancer patients enrolled in a neoadjuvant clinical trial. Transcriptomic analyses demonstrated that AR inhibition drove tumors towards a neuroendocrine-like disease state. In addition, epigenomic profiling revealed massive enzalutamide-induced reprogramming of pioneer factor FOXA1 – from inactive chromatin binding sites towards active *cis*-regulatory elements that dictate pro-survival signals. Notably, treatment-induced FOXA1 sites were enriched for the circadian rhythm core component ARNTL. Post-treatment ARNTL levels associated with poor outcome, and ARNTL suppression decreased cell growth *in vitro*. Our data highlight a remarkable cistromic plasticity of FOXA1 following AR-targeted therapy, and revealed an acquired dependency on circadian regulator ARNTL, a novel candidate therapeutic target.

**Significance:** Understanding how prostate cancer cells adapt to AR-targeted interventions is critical for identifying novel drug targets to improve the clinical management of treatment-resistant disease. Our study revealed an enzalutamide-induced epigenetic plasticity towards pro-survival signaling, and uncovered circadian regulator ARNTL as an acquired vulnerability after AR inhibition, presenting a novel clinical lead for therapeutic development.

## Introduction

Hormonal ablation is the mainstay treatment for patients with metastatic prostate cancer (PCa), ever since the direct critical connection between androgens and prostate tumor progression was first described (1). The androgen receptor (AR) is the key driver of PCa development and progression, and multiple therapeutic strategies have been developed over the years to effectively block the activity of this hormone-driven transcription factor. Upon androgen binding, AR associates with the chromatin at distal *cis*-regulatory enhancer elements, where it regulates the expression of genes through long-range chromatin interactions in three-dimensional genomic space (2, 3). AR does not operate in isolation, but rather recruits a large spectrum of coregulators and other transcription factors to promote expression of genes that drive cancer cell proliferation (4). Critical AR interactors in the transcription complex are HOXB13 and FOXA1, which are both upregulated in primary PCa (4–6) and demarcate enhancers that drive not only primary tumorigenesis but also metastatic disease progression (7). Mechanistically, FOXA1 acts as a pioneer factor, rendering the chromatin accessible for AR to bind (8–11). *FOXA1* is frequently mutated in PCa (12–16) which was shown to alter its pioneering capacities, perturb luminal epithelial differentiation programs, and promote tumor growth, further highlighting the critical role of FOXA1 in human prostate tumors (17, 18). Most patients are diagnosed with organ-confined PCa, which can potentially be cured through locoregional therapies, such as surgery (radical prostatectomy), radiotherapy and/or brachytherapy (19). However, approximately 30% of these patients experience a biochemical recurrence (BCR) – a rise in prostate-specific antigen (PSA) serum levels – indicating PCa relapse (20). At this stage of the disease, suppression of androgen production is a commonly applied therapeutic intervention that can delay further cancer progression for years (21, 22). Nevertheless, the development of resistance to androgen deprivation is inevitable, resulting in castration-resistant prostate cancer (CRPC) for which there is no cure (23). Most CRPC tumors acquired molecular features that enable active AR signaling despite low circulating androgen levels, a finding that led to the development of several highly effective AR-targeted therapies. Enzalutamide (ENZ) is one of the most frequently used AR-targeting agents, which functions through a combined mechanism of blocked AR nuclear import, diminished AR chromatin binding and decreased transcription complex formation, effectively impairing AR-driven PCa growth (24). ENZ’s potent anti-tumor activity has been demonstrated in multiple clinical trials, which led to its FDA approval in various PCa disease stages – from metastatic CRPC (25, 26), to metastatic hormone-sensitive (27), and even non-metastatic CRPC (28) – illustrating how AR-targeted therapies are being progressively introduced earlier in clinical practice.

A clinical benefit of ENZ monotherapy as a neoadjuvant treatment prior to prostatectomy for patients with localized disease, has not been established. Although effective, resistance to AR pathway inhibition will ultimately develop, and the management of advanced PCa with this acquired resistance remains a major clinical challenge, especially since the underlying mechanisms are still not fully elucidated (29). Therefore, furthering our understanding of how ENZ affects PCa biology may lead to the identification of acquired cellular vulnerabilities that could be therapeutically exploited.

To study global drug-induced transcriptional and epigenetic plasticity in human prostate tumors and identify cellular adaptation mechanisms to evade drug treatment, we designed a phase 2 clinical trial to perform multi-omics studies in pre- and post-treatment samples from high-risk localized PCa patients, treated with neoadjuvant ENZ monotherapy. We identified transcriptional reprogramming after treatment, with deactivation of AR signaling and an activation of cell plasticity with neuroendocrine (NE)-like features upon 3 months of AR suppression. Post treatment, these tumors harbored a distinct set of 1,430 *de novo* occupied FOXA1-positive *cis*-regulatory elements, positive for – yet independent of – AR activity, which are dictated by circadian clock core regulator ARNTL to drive tumor cell proliferation instead. Using ARNTL knockdown experiments we could further enhance ENZ sensitivity in cell line models, revealing an unexpected biological interplay between hormonal resistance and circadian rhythm regulation, and identifying a novel highly promising candidate drug target in the clinical management of primary high-risk PCa.

## Results

### Neoadjuvant ENZ therapy for patients with high-risk localized PCa

To study how early ENZ intervention affects prostate tumor biology in a non-castrate environment, we performed integrative multi-omics analyses as part of a single-arm, open-label phase 2 clinical trial: the DARANA study (Dynamics of Androgen Receptor Genomics and Transcriptomics After Neoadjuvant Androgen Ablation; ClinicalTrials.gov number, NCT03297385). In this trial, 56 men with primary high-risk (Gleason score ≥ 7) PCa were enrolled (**Fig. 1A**). Patient demographics and disease characteristics are summarized in **Table 1**, and clinical outcomes of this study are discussed in **Supplementary Data** (**Supplementary Fig. S1**; **Supplementary Table S1)**. Prior to ENZ therapy, magnetic resonance imaging (MRI)-guided core needle tumor biopsies were taken – hereafter referred to as the pre-treatment setting.

**Figure 1:**
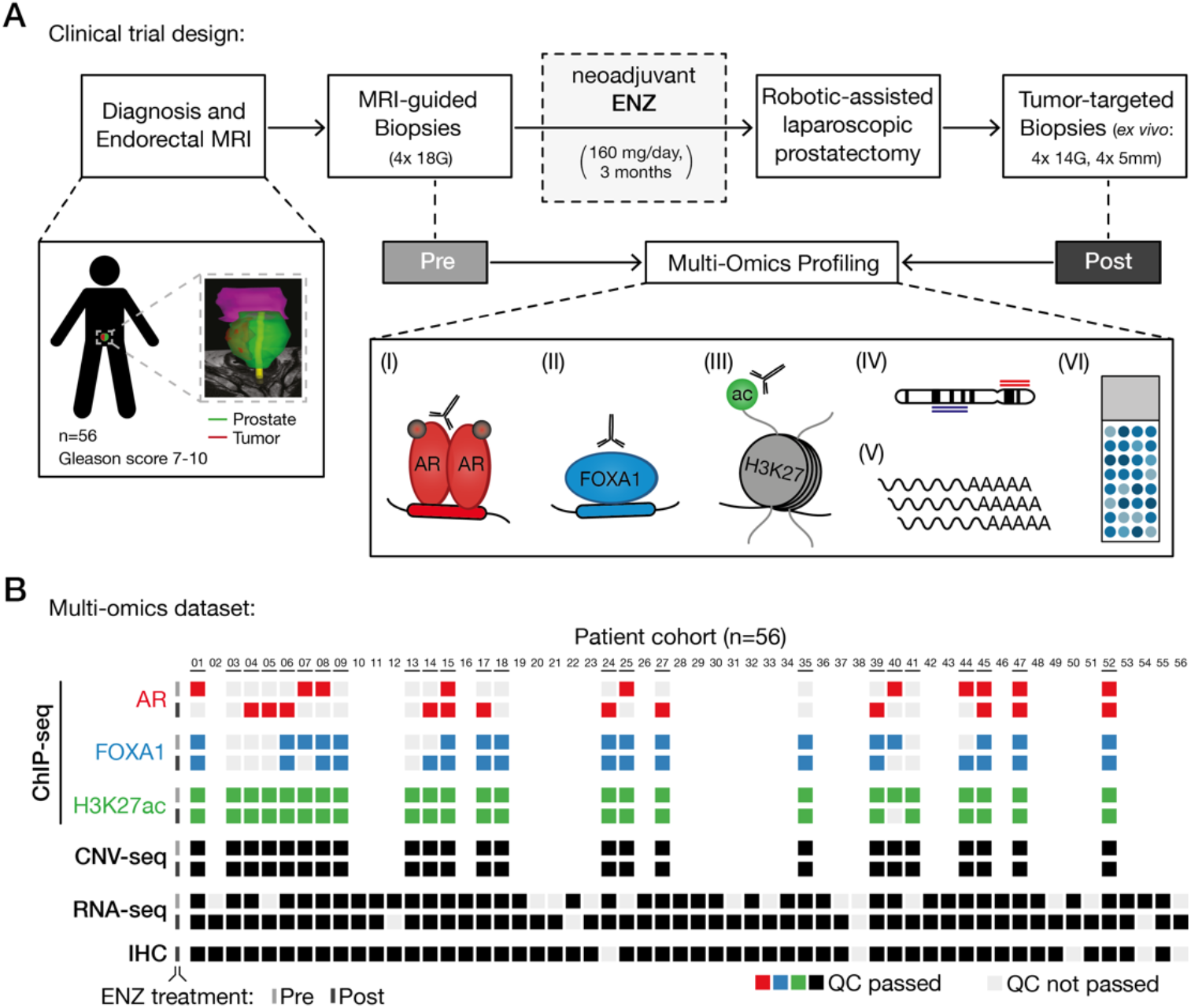
Clinical trial design and omics data sample collection. (A) Study design of the DARANA trial (NCT03297385). Multi-omics profiling, consisting of (I) Androgen Receptor (AR) ChIP- seq, (II) FOXA1 ChIP-seq, (III) H3K27ac ChIP-seq, (IV) DNA copy number sequencing (CNV-seq), (V) gene expression profiling (RNA-seq) and (VI) immunohistochemistry (IHC) analysis, was performed on MRI-guided biopsy samples prior to ENZ treatment (Pre) and tumor-target prostatectomy specimens after 3 months of neoadjuvant ENZ therapy (Post). (B) Overview of data availability and quality control analyses for each sample. Individual data streams are indicated separately with ChIP-seq for AR (red), FOXA1 (blue), H3K27ac (green), CNV-seq, RNA-seq and IHC (all black). The ENZ treatment status indicates the pre-treatment (top) and post-treatment samples (bottom) per omics dataset. Samples not passing QC (light gray) were successfully applied for focused raw data analyses. Blank spots for ChIP-seq or CNV- seq samples indicate that the fresh-frozen material didn’t pass the tumor cell percentage cut-off of ≥ 50%.

**Table 1:**
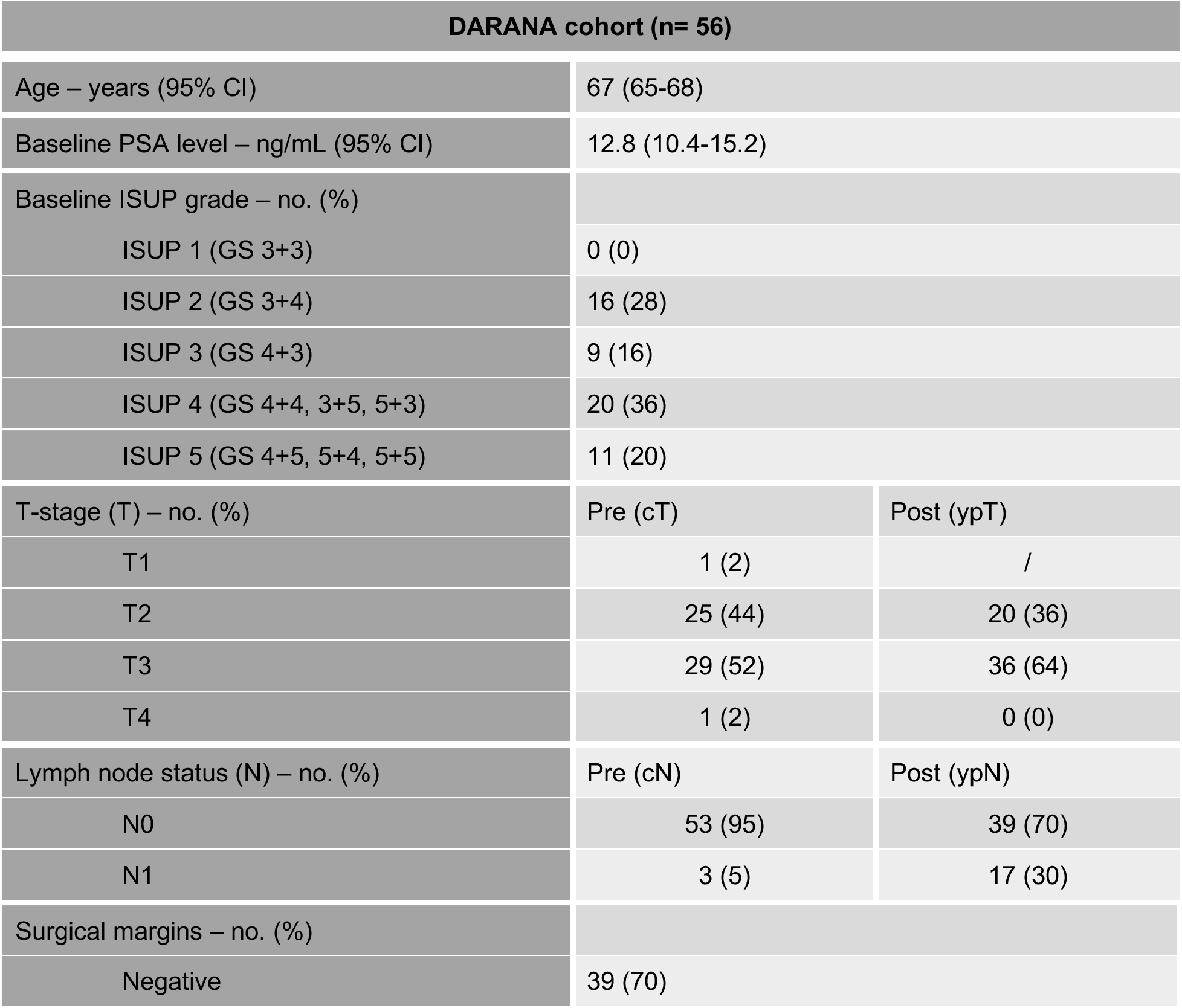
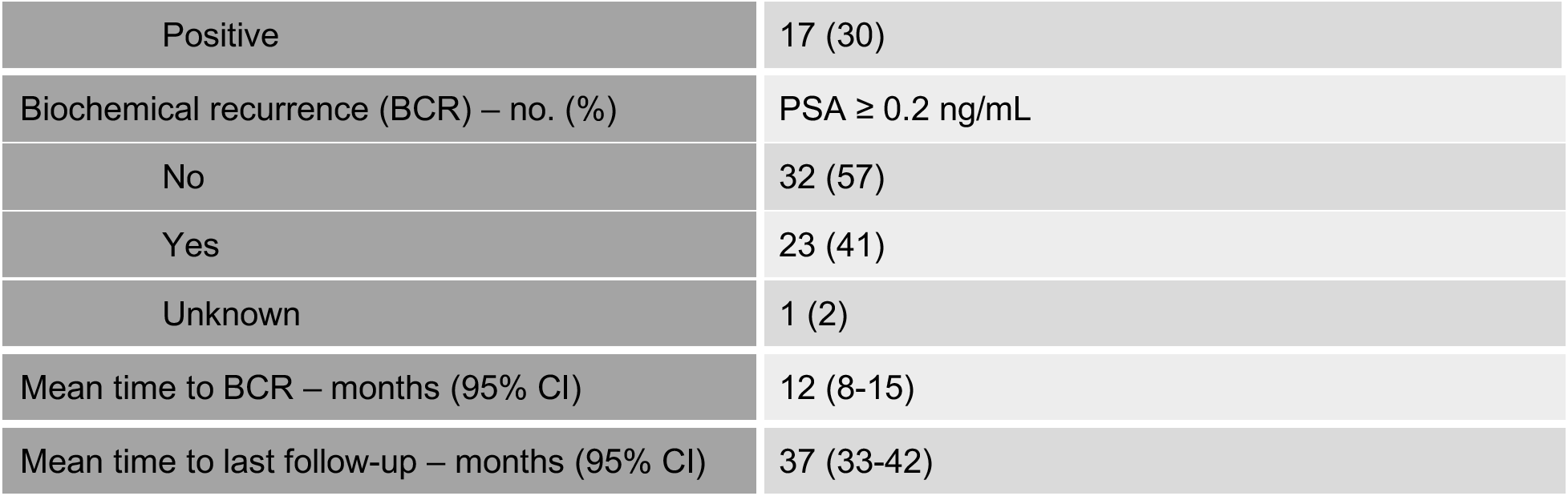
Characteristics of the DARANA cohort (n=56). Table summarizing the patient baseline demographics, and pre- and post-treatment disease characteristics of the DARANA cohort. Shown are age (years), initial Prostate-specific antigen (PSA) serum levels (ng/mL) and International Society of Urological Pathology (ISUP) grade at diagnosis (with associated Gleason scores (GS)). In addition, T-stage (T) and Lymph node status (N) before (pre = at diagnosis) and after (post = at surgery) neoadjuvant ENZ therapy, as well as the surgical margin status of the prostatectomy specimens are shown. Pre-treatment measures are based on histological evaluation of biopsy material and radiographic evaluation (clinical grading; c), while post-treatment assessments are based on histological evaluations of prostatectomy specimens (pathological grading after neoadjuvant therapy; yp). Biochemical recurrence (BCR) was defined as a rise in PSA of ≥ 0.2 ng/mL. Mean time to BCR (months) and time to last follow-up (months) are indicated. For continuous variables (age, baseline PSA, time to BCR, and time to last follow-up) the mean and 95% confidence interval (CI) are shown. For categorical variables (baseline ISUP, T-stage, N-status, surgical margins, BCR) the number of patients (no.) and percentages (%) are indicated.

| DARANA cohort (n= 56) |  |  |
| --- | --- | --- |
| Age – years (95% CI) | 67 (65-68) |  |
| Baseline PSA level – ng/mL (95% CI) | 12.8 (10.4-15.2) |  |
| Baseline ISUP grade – no. (%) |  |  |
| ISUP 1 (GS 3+3) | 0 (0) |  |
| ISUP 2 (GS 3+4) | 16 (28) |  |
| ISUP 3 (GS 4+3) | 9 (16) |  |
| ISUP 4 (GS 4+4, 3+5, 5+3) | 20 (36) |  |
| ISUP 5 (GS 4+5, 5+4, 5+5) | 11 (20) |  |
| T-stage (T) – no. (%) | Pre (cT) | Post (ypT) |
| T1 | 1 (2) | / |
| T2 | 25 (44) | 20 (36) |
| T3 | 29 (52) | 36 (64) |
| T4 | 1 (2) | 0 (0) |
| Lymph node status (N) – no. (%) | Pre (cN) | Post (ypN) |
| N0 | 53 (95) | 39 (70) |
| N1 | 3 (5) | 17 (30) |
| Surgical margins – no. (%) |  |  |
| Negative | 39 (70) |  |

|  |  |
| --- | --- |
| Positive | 17 (30) |
| Biochemical recurrence (BCR) – no. (%) | PSA ≥ 0.2 ng/mL |
| No | 32 (57) |
| Yes | 23 (41) |
| Unknown | 1 (2) |
| Mean time to BCR – months (95% CI) | 12 (8-15) |
| Mean time to last follow-up – months (95% CI) | 37 (33-42) |

Subsequently, patients received neoadjuvant ENZ treatment (160 mg/day) without androgen deprivation therapy for three months, followed by robotic-assisted laparoscopic prostatectomy. Based on baseline MRI information and palpation, additional tumor-targeted core needle biopsies were taken *ex vivo* – representing the post-treatment setting. This pre- and post-treatment sampling allowed us to study the epigenetic, genomic, transcriptomic and proteomic effects of neoadjuvant ENZ therapy in individual patients (**Fig. 1A**). We generated chromatin immunoprecipitation (ChIP-seq) profiles of the prostate cancer drivers AR and FOXA1, as well as the histone modification H3K27ac before and after ENZ treatment, and integrated these cistromic findings with pre- and post-treatment gene expression (RNA-seq), copy number (CNV-seq) and immunohistochemistry (IHC) data from the same tumors. Stringent quality control (QC) analyses were performed on all data streams (**Supplementary Fig. S1B**), and the following number of samples passed all QC measures (**Fig. 1B**): AR ChIP-seq (pre: n=10; post: n=12), FOXA1 ChIP-seq (pre: n=17; post: n=17), H3K27ac ChIP-seq (pre: n=24; post: n=23), CNV-seq (pre: n=24; post: n=24), RNA-seq (pre: n=42; post: n=52) and IHC (post: n=51).

Collectively, we performed integrative multi-omics analyses as part of a clinical trial that enabled us to examine ENZ-induced oncogenomic changes to identify early epigenetic steps in treatment response, but also therapy-induced resistance.

### Characterization of tissue ChIP-seq data

To assess how neoadjuvant ENZ treatment affects the *cis*-regulatory landscape in primary PCa, we generated human tumor ChIP-seq profiles for the transcription factors AR and FOXA1, along with the active enhancer/promoter histone mark H3K27ac before and after neoadjuvant intervention. ChIP-seq quality metrics are summarized in **Supplementary Fig. S2** and **Supplementary Table S2**. Visual inspection at known AR target genes showed high-quality data for all ChIP-factors in both clinical settings (**Fig. 2A**). On a genome-wide scale, the H3K27ac ChIP-seq profiles were highly distinct from the transcription factors (TFs) and divided the samples into two main clusters irrespective of their treatment status (**Fig. 2B and 2C**). Notably, AR and FOXA1 ChIP-seq datasets were intermingled in the clustering analysis, suggesting largely comparable binding profiles which is in line with FOXA1’s role as a canonical AR pioneer factor (**Supplementary Fig. S3**) (5, 30). As described previously (31), highest Pearson correlation was found between H3K27ac samples, indicating comparable histone acetylation profiles among primary PCa samples (**Fig. 2B**; **Supplementary Fig. S3**). Much greater heterogeneity in chromatin binding was observed for the TFs AR and FOXA1, which is further supported by the steep decrease in the number of overlapping AR and FOXA1 peaks with increasing number of samples compared to H3K27ac (**Fig. 2D**; **Supplementary Fig. S3**). In order to maintain the high-confidence peaks that have been reproducibly identified in multiple patients without losing too much binding site heterogeneity between samples, we decided to generate consensus peaksets. To this end, we only considered binding sites that were present in at least 3 out of 22 AR samples, 7 out of 34 FOXA1 samples and 13 out of 47 H3K27ac samples, which corresponds to ∼25% of all binding sites identified for each factor (**Fig. 2D**). Genomic distribution analyses of these consensus sites revealed distinct enrichments for annotated genomic regions: While AR and FOXA1 were almost exclusively found at intronic and distal intergenic regions, H3K27ac peaks were also enriched at promoters (**Fig. 2E**), which is in line with previously published genomic distributions of AR (5, 31), FOXA1 (5, 9), and H3K27ac (31, 32). In addition, motif enrichment analyses at AR and FOXA1 consensus peaks identified, as expected, androgen and Forkhead response elements among the top-ranked motifs, respectively (**Fig. 2F**).

**Figure 2:**
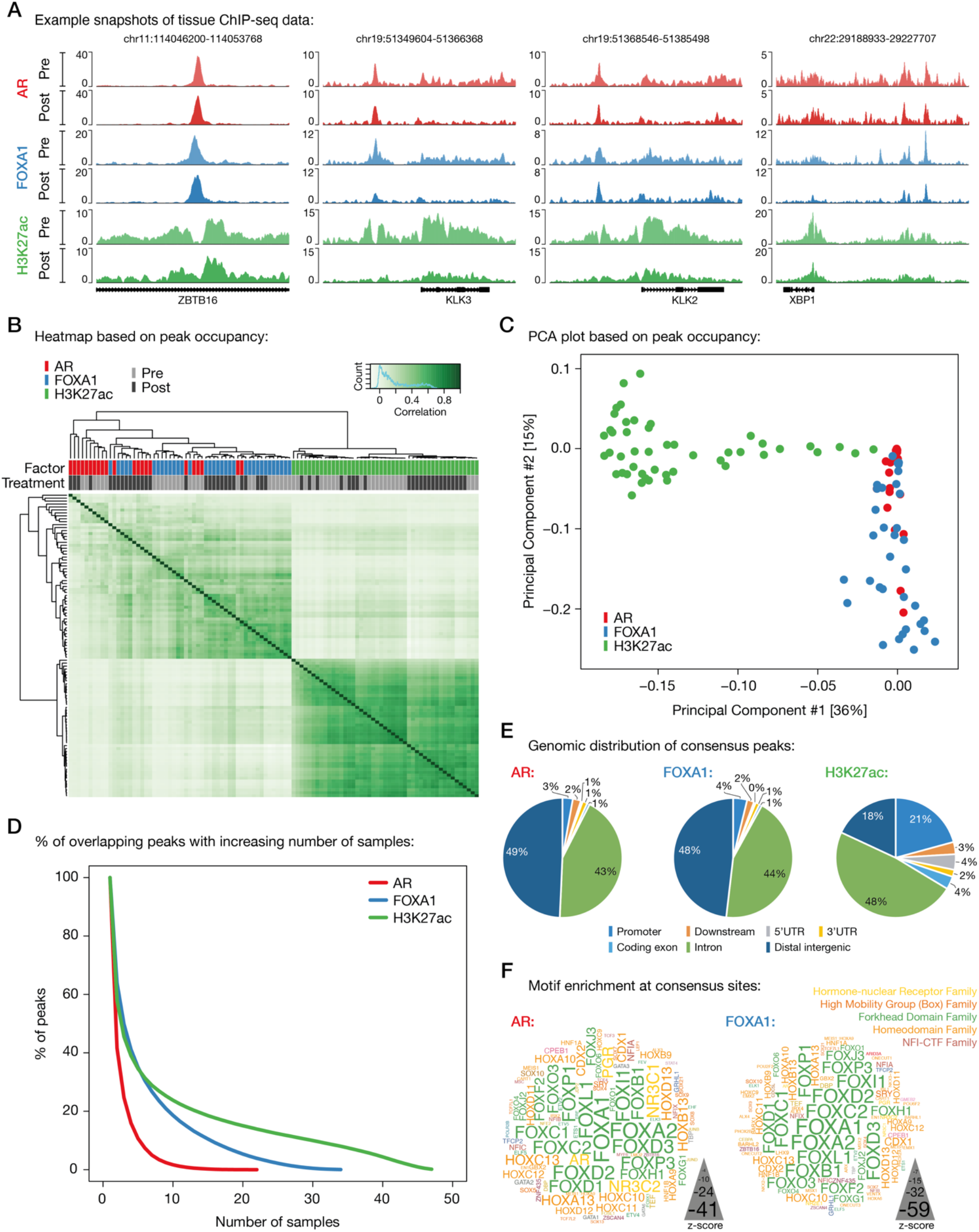
Characterization of tissue ChIP-seq data streams. (A) Representative example snapshots of AR (red), FOXA1 (blue) and H3K27ac (green) ChIP-seq data for four genomic loci in one patient. Pre- (light colors) and post-ENZ treatment (dark colors) is indicated. Y-axis indicates ChIP-seq signal in fragments per kilobase per million reads mapped (FPKM). (B) Correlation heatmap based on peak occupancy. Clustering of the samples is based on all called peaks and represents Pearson correlations between individual ChIP-seq samples. The column color bars indicate the ChIP-seq factor (AR, FOXA1, H3K27ac) and treatment status (Pre, Post). (C) Principal component analysis (PCA) plot based on peak occupancy. Each dot represents a ChIP-seq sample that is colored per factor. (D) Elbow plot depicting the peak overlap between ChIP-seq samples per factor. Shown is the percentage of overlapping peaks with increasing number of samples. Consensus peaksets were designed by using a cut-off of peaks present in at least 3 AR, 7 FOXA1, or 13 H3K27ac samples. (E) Pie charts showing the genomic distribution of AR (left), FOXA1 (middle) and H3K27ac (right) consensus peaks. (F) Word clouds show motif enrichment at AR (left) and FOXA1 (right) consensus sites. The font size represents the z- score and colors correspond to transcription factor families.

Taken together, we generated multiple high-quality tissue ChIP-seq data streams that now allowed us to study ENZ-induced changes in primary PCa patients.

### ENZ treatment enriches for newly acquired FOXA1-bound regulatory regions

To identify ENZ-induced TF reprogramming and epigenetic changes, we performed differential binding analyses comparing the pre- and post-treatment tissue ChIP-seq samples. Therefore, we first ran occupancy-based unsupervised principal component analyses (PCA) to detect whether ENZ treatment led to differences in TF chromatin binding. While the sample size of the AR ChIP-seq data stream was not sufficient to observe significant differences in peak occupancy pre- versus post-treatment (**Supplementary Fig. S4A**), the FOXA1 data did show such differences, with a clear separation of pre- and post-treatment FOXA1 samples in the second principal component (**Fig. 3A**). Subsequent supervised analysis (pre vs. post) revealed a total of 1,905 genomic regions (475 pre-enriched, 1,430 post-enriched; **Supplementary Table S3**) that showed significant differential FOXA1 binding between both clinical settings (FDR < 0.05; **Fig. 3B and 3C**; **Supplementary Fig. S4B and S4C**). Further characterization of these differential FOXA1 regions showed that both sets of binding sites were still preferentially located in intronic and distal intergenic regions (with a slight enrichment for promoters at the post-enriched sites; **Supplementary Fig. S4D**). In addition, Forkhead domain family motifs were the top enriched motifs at both pre- and post-enriched sites, illustrating that treatment does not alter FOXA1 motif preference and still occupies canonical FOXA1 binding sites (**Supplementary Fig. S4E**).

**Figure 3:**
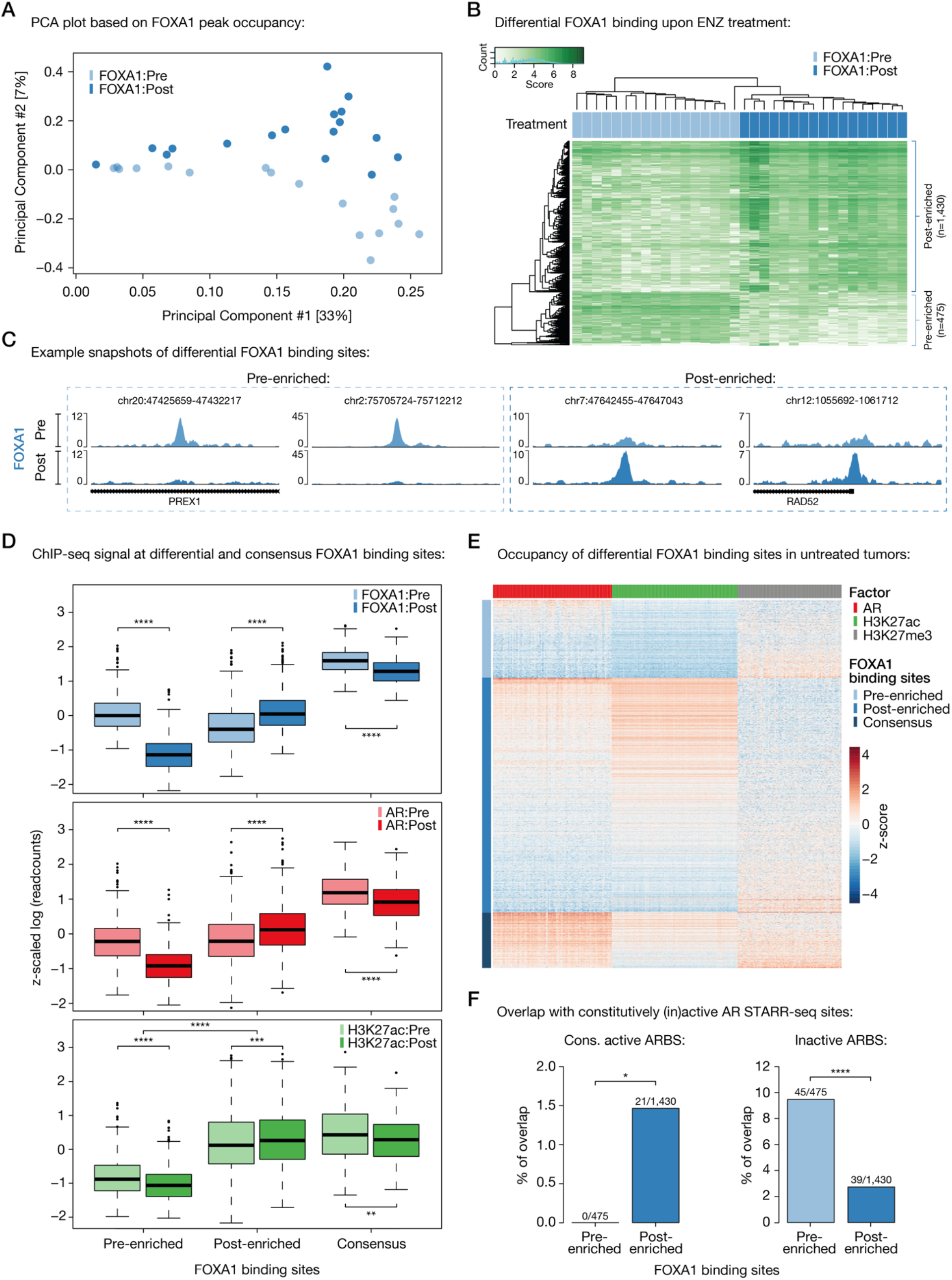
Differential FOXA1 binding upon ENZ treatment. (A) Principle component analysis (PCA) plot based on peak occupancy of FOXA1 ChIP-seq data. Color indicates pre-treatment (light blue) and post-treatment (dark blue) FOXA1 samples. (B) Coverage heatmap depicting differential FOXA1 binding sites, selectively enriched in the pre-treatment (n=475) or post-treatment (n=1,430) setting. (C) Representative example snapshots of FOXA1 ChIP-seq signal at two pre-enriched (left) and two post-enriched (right) FOXA1 sites in one patient. Pre- (light blue) and post-ENZ treatment (dark blue) is indicated. Y-axis indicates ChIP-seq signal in FPKM. (D) Boxplots indicating ChIP-seq signal (z-scaled readcounts) at pre-enriched (n=475), post-enriched (n=1,430) and consensus FOXA1 peaks (shared by ≥ 30 patients; n=338) for FOXA1 (blue), AR (red), and H3K27ac (green) ChIP-seq datasets before (Pre; light colors) and after (Post; dark colors) ENZ treatment. **, *P* < 0.01; ***, *P* < 0.001; ****, *P* < 0.0001 (Mann-Whitney U-test adjusted for multiple testing using FDR). (E) Coverage heatmap showing occupancy of differential (pre-/post-enriched) and consensus FOXA1 peaks in an external ChIP-seq dataset consisting of 100 untreated primary tumors (31). Heatmap color indicates region read counts (z-score) at pre-enriched, post-enriched and consensus FOXA1 sites (rows) in the AR (red), H3K27ac (green) and H3K27me3 (gray) ChIP-seq data streams (columns). (F) Bar chart representing the overlap between differential FOXA1 sites (pre-enriched or post-enriched) and constitutively active (left) or inactive (right) AR binding sites (ARBS), based on STARR-seq. *, *P* < 0.05; ****, *P* < 0.0001 (Fisher’s exact test).

To examine whether structural variations are underlying these differential FOXA1 binding events, we performed CNV-seq on the same tumor specimens and then projected onto the differential FOXA1 cistromics the structural copy-number data. These analyses revealed a comparable level of CNV at pre- and post-treatment enriched FOXA1 sites before and after ENZ treatment, with an overall trend towards less CNV upon treatment (**Supplementary Fig. S5A-S5C)**. However, in none of the matched sample pairs (pre and post CNV-seq, and FOXA1 ChIP-seq; n=15) a strong correlation between copy number difference and ChIP-seq signal difference was observed (*R* = 0.11; **Supplementary Fig. S5D**). In total, at only 44 out of 1,905 differential FOXA1 binding sites (< 2.5%), we observed copy number differences between post- and pre-treatment samples that could potentially explain binding site occupancy in 3 or more patients, indicating that the vast majority of these differential binding events is based on treatment-induced transcription factor reprogramming, rather than structural variation (**Supplementary Fig. S5E**).

As FOXA1 dictates AR chromatin binding capacity (5), epigenetic plasticity of FOXA1 induced by treatment may be associated with alterations in the AR cistrome. To assess this, and to explore the epigenetic landscape surrounding the differentially bound FOXA1 regions, we compared the ChIP-seq signal of all three factors (AR, FOXA1, H3K27ac) at differential (pre- / post-enriched) and consensus (shared by ≥ 30 patients; n=338) FOXA1 sites before and after ENZ therapy. While the FOXA1 ChIP-seq signal was highest at consensus binding sites, the pre- and post-treatment enriched regions followed the expected trend and showed significantly higher signal in the corresponding settings (**Fig. 3D**). Notably, we also observed less binding of FOXA1 to consensus sites when treated with ENZ, although the differences are much milder compared to the effects seen at pre-enriched FOXA1 sites (*Padj* = 3.62 x 10^-22^ at consensus vs. 3.76 x 10^- 130^ at pre-enriched sites, Mann-Whitney U-test; **Fig. 3D**; **Supplementary Fig. S6A**). This could possibly be explained by decreased FOXA1 gene expression levels upon ENZ treatment (**Supplementary Fig. S6B**).

The AR ChIP-seq signal followed the same patterns as observed for FOXA1, suggesting that relocated FOXA1 upon treatment functionally drives alterations in the AR cistrome (**Fig. 3D**). Unexpectedly, the pre-enriched FOXA1 sites were completely devoid of any H3K27ac signal in both pre- and post-treatment samples, while the post-enriched counterparts were positive for this active enhancer/promoter mark with a significant increase post-ENZ (*Padj* = 5.59 x 10^-4^, Mann-Whitney U-test; **Fig. 3D**; **Supplementary Fig. S6C and S6D**), suggesting that pre-ENZ FOXA1 sites are inactive. To validate these observations in an independent cohort, we analyzed previously published AR (n=87), H3K27ac (n=92) and H3K27me3 (n=76) ChIP-seq data from a cohort of 100 primary treatment-naïve PCa samples (31). Supporting our previous analyses, the vast majority of post-enriched FOXA1 sites were H3K27ac-positive and their histone acetylation status positively correlated with AR binding (*R* = 0.78) (**Fig. 3E**; **Supplementary Fig. S6E**). The pre-enriched FOXA1 sites, however, were again H3K27ac-negative, while the repressive histone modification H3K27me3 was present, which further points towards an inactive epigenetic state of these regulatory regions (**Fig. 3E**).

Recently, we reported that prostate cancers can reactivate developmental programs during metastatic progression (7). These sentinel enhancers appeared to be premarked by FOXA1 from prostate gland development, and albeit inactive in normal and primary tumor specimens, the sites get reactivated by AR during metastatic outgrowth. Given the inactivity of the pre-enriched FOXA1 sites, we hypothesized that FOXA1 might be decommissioned at such developmental enhancers prior to hormonal intervention. To test this, we overlapped the differential FOXA1 binding sites with the metastasis-specific AR binding sites (met- ARBS; n=17,655), which revealed a strong enrichment for these developmental regulatory elements at pre-treatment FOXA1 sites (*P* = 2.13 x 10^-16^, Fisher’s exact test; **Supplementary Fig. S6F**). But are the inactive pre-enriched FOXA1 sites solely epigenetically suppressed, or are these regions intrinsically incapable of being active in this cellular context? To address this question and to further elucidate the role of AR at these differentially bound FOXA1 sites, we integrated our tissue ChIP-seq findings with previously identified tumor-specific AR binding sites (n=3,230) (5) that were functionally characterized using Self-Transcribing Active Regulatory Regions sequencing (STARR-seq), a massive parallel reporter assay to systematically annotate intrinsic enhancer activity (33). With this, three distinct classes of AR binding sites (ARBS) were identified (**Supplementary Table S4)**: enhancers that were active regardless of AR stimulation (constitutively active; n=465), ARBS with no significant enhancer activity (inactive; n=2,479) and inducible AR enhancers that increase activity upon androgen treatment (inducible; n=286). Interestingly, we found that post-treatment FOXA1 sites were enriched for constitutively active ARBS, which further supports the high enhancer activity and H3K27ac positivity observed at these sites, but also illustrates that this activity is constitutive and AR-independent (**Fig. 3F**). Consistent with our postulated inactivity of the pre-treatment enriched FOXA1 sites, these regions overlapped highly significantly with inactive ARBS (*P* = 8.60 x 10^-9^, Fisher’s exact test), which implies that these DNA elements are intrinsically inactive and incapable to act as functional enhancers, and possibly explains why these AR-bound sites did not show active regulatory marks (**Fig. 3E and 3F**). As no enrichment of our differential FOXA1 sites was observed with inducible ARBS (Pre-enriched: 4/475; Post-enriched: 2/1,430), these data further support a conclusion that AR itself is not a driver at FOXA1 sites that are differentially occupied after ENZ exposure in patients.

Overall, these results suggest that prior to hormonal intervention, FOXA1 is decommissioned at inactive developmental enhancer elements, which based on their primary DNA sequence are intrinsically incapable of being active – at least in the tested hormone-sensitive disease setting. However, upon ENZ treatment, FOXA1 gets reprogrammed to highly active *cis*-regulatory regions, which act in an AR-independent manner.

### Transcriptional rewiring upon neoadjuvant ENZ

Having assessed the cistromic and epigenomic changes in response to neoadjuvant ENZ, we next determined how transcriptional programs were affected by this hormonal intervention. Principal component analysis (PCA) across both treatment states revealed that three months of ENZ therapy has a major effect on global gene expression profiles (**Fig. 4A**). Subsequently, we performed differential gene expression analysis, in which we compared pre- and post-treatment RNA-seq samples. Gene set enrichment analysis (GSEA) showed that AR signaling, along with mitosis and MYC signals, was strongly decreased upon treatment (**Fig. 4B and C**; **Supplementary Fig. S7A**). Since ENZ blocks the AR signaling axis, we analyzed the androgen-response pathway in more detail, which revealed a strong downregulation of AR target genes in almost every patient (**Fig. 4D**). In contrast to this, TNF*α* signaling, IFN-*γ* response and epithelial-mesenchymal transition (EMT) signals were most upregulated (**Fig. 4B**; **Supplementary Fig. S7B**).

**Figure 4:**
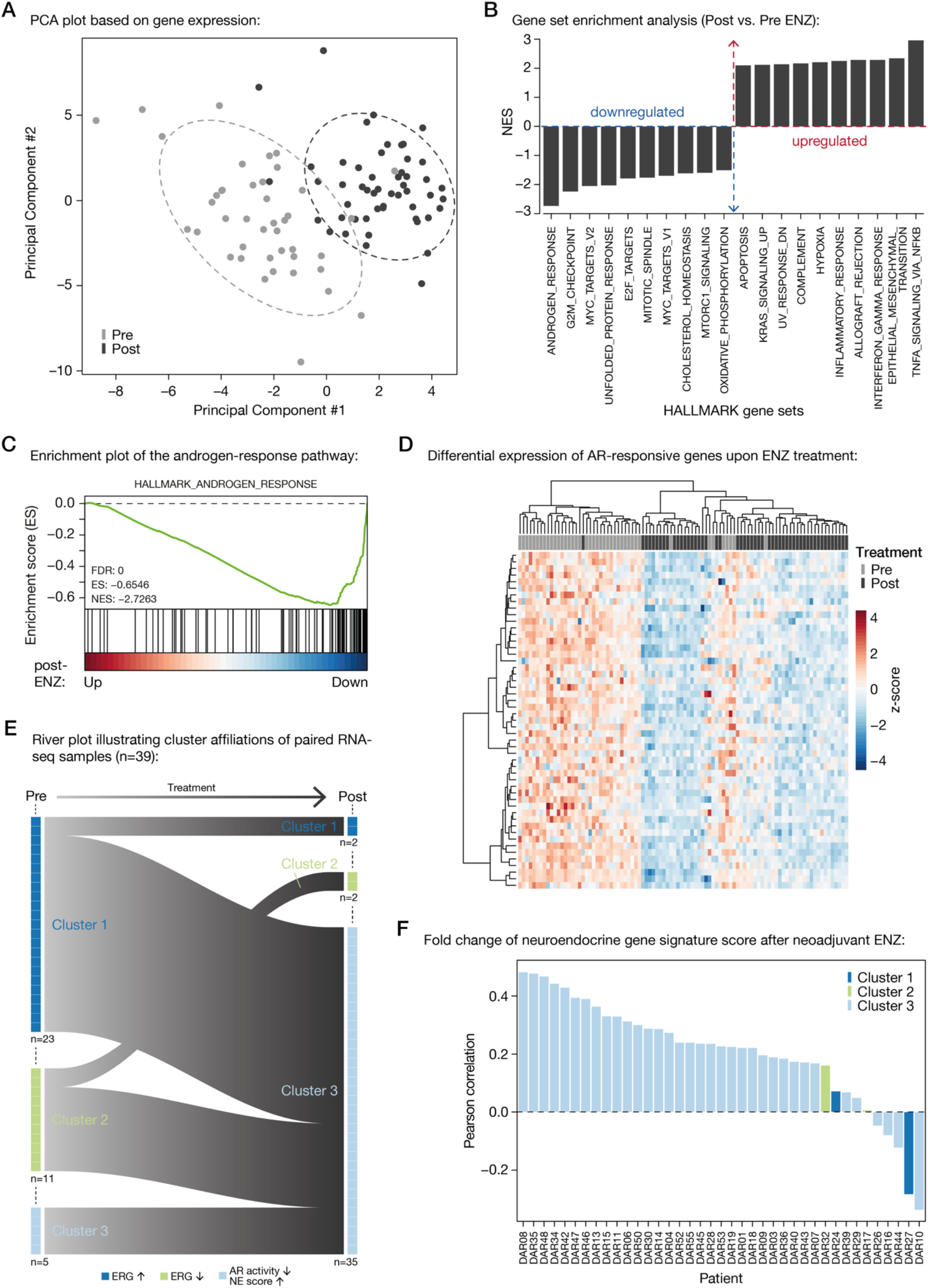
Neoadjuvant ENZ deactivates AR signaling and induces neuroendocrine (NE)-like gene expression signatures. (A) Principle component analysis (PCA) plot based on gene expression data. Color indicates pre-treatment (gray) and post- treatment (black) samples. Ellipses are based on the 80% confidence interval. (B) Gene set enrichment analyses (GSEA) for Hallmark gene sets. Shown are the top differentially enriched pathways upon ENZ treatment. Y-axis indicates the normalized enrichment score (NES). (C) Enrichment plot of the Hallmark Androgen Response pathway, for genes up- or downregulated after ENZ. Y-axis indicates enrichment score (ES). GSEA statistics (FDR, ES, NES) are indicated. (D) Unsupervised hierarchical clustering of pre- and post-treatment RNA-seq samples based on the expression of AR- responsive genes. Color scale indicates gene expression (z-score). (E) River plot showing state transitions between Clusters 1 (dark blue), Cluster 2 (green) and Cluster 3 (light blue) for paired pre-treatment and post-treatment RNA-seq samples (n=39). Number of samples assigned to each cluster before and after treatment as well as the hallmarks per cluster are indicated. (F) Waterfall plot depicting the Pearson correlation of neuroendocrine gene expression signature fold changes upon ENZ treatment per patient. Colors indicate the patients cluster affiliations after treatment.

Previously, we identified three distinct subtypes of primary treatment-naïve PCa (31), which we named Cluster 1-3 (Cl1-3). While Cl1 and Cl2 were mainly dominated by their ERG fusion status – with Cl1 expressing high ERG levels (ERG fusion-positive) and Cl2 expressing low ERG levels (ERG fusion-negative) – Cl3 was enriched for neuroendocrine (NE)-like features, including low AR activity and a high NE gene expression score. To assess the impact of neoadjuvant ENZ therapy on these PCa subtypes, we performed unsupervised hierarchical clustering in the pre- and post-treatment setting using the originally identified top 100 most differentially expressed genes per cluster. Prior to hormonal intervention, we could robustly assign the samples into all three clusters (Cl 1: n=23, Cl2: n=11, Cl3: n=8) with highly comparable distributions as we previously reported in another cohort of patients (31) (**Supplementary Fig. S8A**). Our pre- and post-treatment sampling now allowed us to investigate how individual tumors were affected by neoadjuvant therapy. This revealed that three months of ENZ therapy pushed almost all of the tumors towards our NE-like cluster 3 (**Fig. 4E**; **Supplementary Fig. S8B**). To assure that the observed effects are not solely driven by the treatment-induced reduction in AR activity (**Fig. 4C and 4D**), we used a well-established neuroendocrine PCa (NEPC) signature (34) to calculate gene expression fold changes pre- vs. post-ENZ, which confirmed an induction of NE-like signaling upon treatment (**Fig. 4F**).

Collectively, these results demonstrate that three months of neoadjuvant ENZ therapy not only uniformly diminishes AR signaling, but also pushes practically all of our primary PCa samples towards a NE-like gene expression state independently of their original subtype.

### Post-treatment FOXA1 sites drive pro-survival gene programs, dictated by circadian clock component ARNTL

Having examined the global cistromic and transcriptomic changes upon ENZ therapy, we next characterized the biological consequences of the observed FOXA1 reprogramming using integrative analyses. We hypothesized that the newly acquired FOXA1 sites would be driving expression of genes associated with tumor cell survival programs. Using H3K27ac HiChIP data generated in LNCaP cells (35), pre- and post-treatment FOXA1 sites were coupled to their corresponding gene promoters (**Supplementary Table S5**). Subsequently, genome-wide CRISPR knockout screen data from Project Achilles (DepMap 20Q1 Public; VCaP) were used to identify those genes essential for prostate cancer cell proliferation (36, 37). While genes associated with pre-treatment FOXA1 sites were not enriched for essentiality as compared to all other CRISPR-targeted genes in the library, genes under control of post-treatment FOXA1 sites showed a significant enrichment (*P* = 7.54 x 10^-12^, Welch’s t-test) for critical drivers of tumor cell proliferation (**Fig. 5A**).

**Figure 5:**
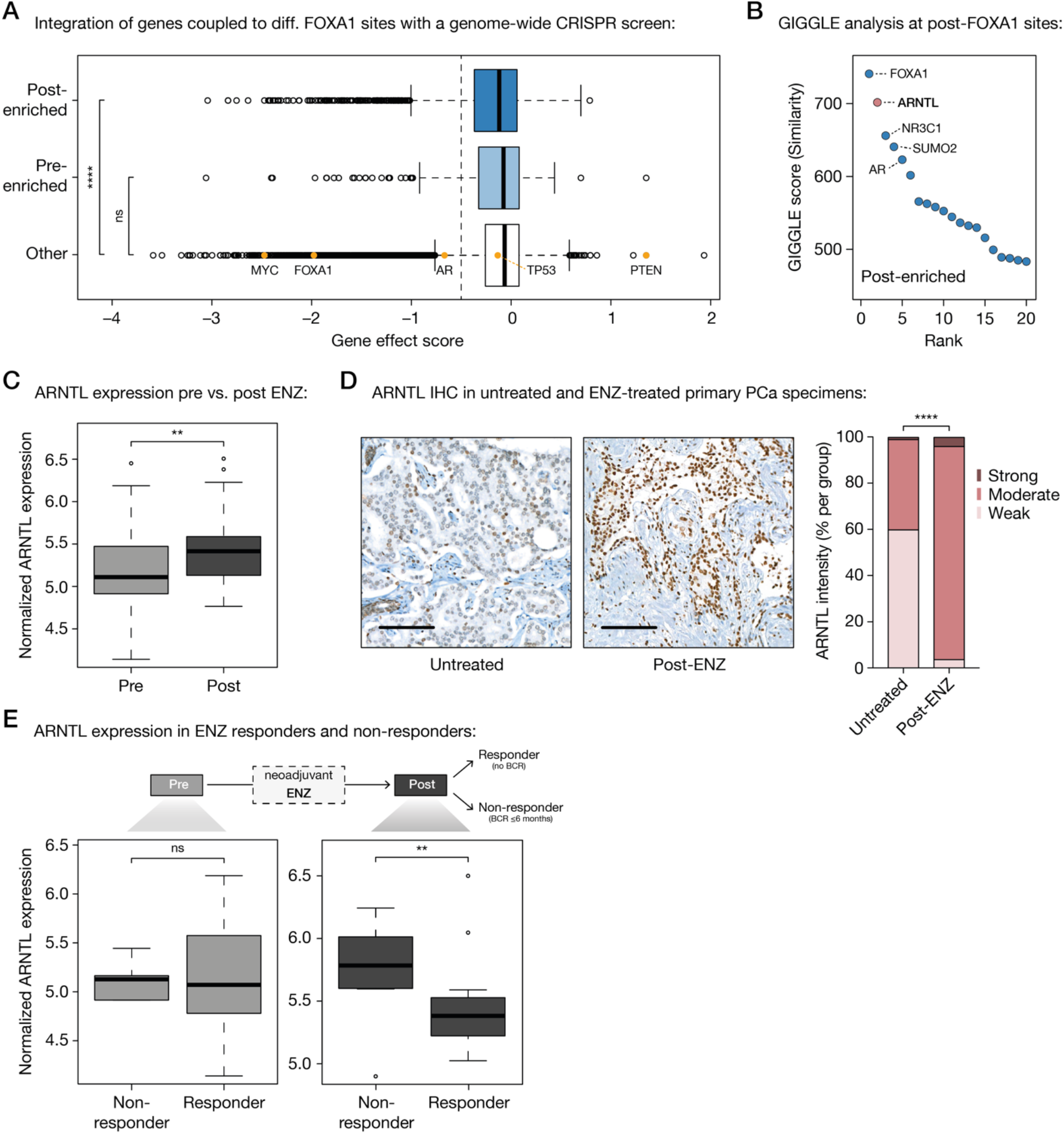
Acquired FOXA1 sites drive key-survival genes that are under control of circadian rhythm regulator ARNTL. (A) Boxplot showing DepMap (20Q1) genome-wide loss-of-function CRISPR screen data for VCaP PCa cells, separately analyzing the gene effect score of genes associated with post-enriched FOXA1 sites (top), pre-enriched FOXA1 sites (middle) or all other tested genes (bottom). Differential FOXA1 binding sites were coupled to their respective target genes using H3K27ac HiChIP data. Indicated as controls are PCa-relevant driver genes: oncogenes *MYC*, *FOXA1*, *AR*, *TP53*and tumor suppressor *PTEN*. The recommended gene effect score cutoff of -0.5 is shown (dotted vertical line). ns, *P* > 0.05; ****, *P* < 0.0001 (Fisher’s exact test). (B) Dot plot representing ranked GIGGLE similarity scores for transcriptional regulators identified at post-treatment FOXA1 sites. The top 20 factors identified are shown, and the 5 most enriched factors are labeled. (C) Boxplot showing normalized ARNTL gene expression before and after 3 months of neoadjuvant ENZ treatment. **, *P* < 0.01 (Mann-Whitney U-test). (D) Representative ARNTL immunohistochemistry (IHC) stainings (left) and quantification of ARNTL staining intensity (right) in tissue microarrays consisting of prostatectomy specimens from untreated patients (not receiving neoadjuvant ENZ; n=110) and DARANA patients post-ENZ (n=51). Scale bars, 100 µm. ****, *P* < 0.0001 (Fisher’s exact test). (E) Boxplots depicting normalized ARNTL gene expression in ENZ non-responders (biochemical recurrence (BCR) ≤ 6 months; n=8) and responders (no BCR; n=29) in the pre- (left) and post- (right) treatment setting separately. ns, *P* > 0.05; **, *P* < 0.01 (Mann-Whitney U-test).

Based on our STARR-seq and RNA-seq data, we concluded that AR is likely not driving enhancer activity at post-treatment FOXA1 sites (**Fig. 3F**; **Fig. 4C and 4D**). Therefore, we sought to identify transcription factors involved in the activation of these regulatory regions that are selectively occupied by FOXA1 following treatment. To this end, we overlaid the genomic coordinates of the post-treatment enriched FOXA1 binding sites with those identified in publicly available ChIP-seq datasets (n= 13,976) as part of the Cistrome DB transcription factor ChIP-seq sample collection (38, 39). Besides FOXA1 and AR, which were expected to bind at these regions (**Fig. 3D**), we also identified the glucocorticoid receptor (encoded by the *NR3C1* gene), which has previously been described to be upregulated upon antiandrogen treatment and able to drive the expression of a subset of AR-responsive genes, conferring resistance to AR blockade (40–42). Unexpectedly, the second most enriched transcription factor after FOXA1 was circadian rhythm core component ARNTL (Aryl Hydrocarbon Receptor Nuclear Translocator Like; also known as BMAL1) which has not previously been implicated in PCa biology (**Fig. 5B**). Interestingly, ARNTL transcript levels were upregulated upon ENZ treatment (*P* = 6.4 x 10^-3^, Mann-Whitney U-test; **Fig. 5C**), which was accompanied by increased H3K27ac ChIP-seq signals at the *ARNTL* locus (**Supplementary Fig. S9A**). Consistent with this, tissue microarray IHC analysis also revealed elevated ARNTL protein levels after treatment when comparing the prostatectomy specimens post-ENZ with those of matched untreated control patients (*P* = 6.89 x 10^-19^, Fisher’s exact test; **Fig. 5D**). To assess whether ARNTL levels are also associated with patient outcome, we compared the average ARNTL gene expression of patients that did not experience a BCR (responders, n=29) with those that experienced an early BCR within ≤ 6 months post-surgery (non-responders, n=8; **Supplementary Table S1**). While pre-treatment ARNTL levels were not significantly different between ENZ responders and non-responders, high ARNTL levels after treatment were associated with poor clinical outcome (*P* = 4.79 x 10^-3^, Mann-Whitney U-test; **Fig. 5E**). Notably, the CLOCK protein, which forms a heterodimer with ARNTL to activate transcription of core clock genes, didn’t show differential expression upon ENZ treatment (**Supplementary Fig. S9B**) and is also not associated with clinical outcome (**Supplementary Fig. S9C**), hinting towards a treatment-induced role of ARNTL that is independent of its canonical function in the circadian machinery.

Taken together, these data suggest that the circadian clock regulator ARNTL may be functionally involved in ENZ resistance by driving tumor cell proliferation processes.

### Acquired ARNTL dependency in ENZ-resistant PCa cells

To further investigate the relevance of ARNTL as a transcriptional driver at post-treatment FOXA1 sites, we performed *in vitro* validation experiments. To this end, we used hormone-sensitive LNCaP PCa cells, which we either cultured in full medium alone (Pre^LNCaP^) or with ENZ for 48 hours (Post^LNCaP^), mimicking our clinical trial setting (**Fig. 6A**). Based on the acquisition of NE-like gene expression profiles post-ENZ (**Fig. 4E and 4F**), we also included the ENZ-resistant LNCaP-42D model (43) that possesses NE-features (Res^LNCaP-42D^; **Fig. 6A**), allowing us to further validate our patient-derived findings in cell lines recapitulating the transcriptional features of post-treatment clinical specimens.

**Figure 6:**
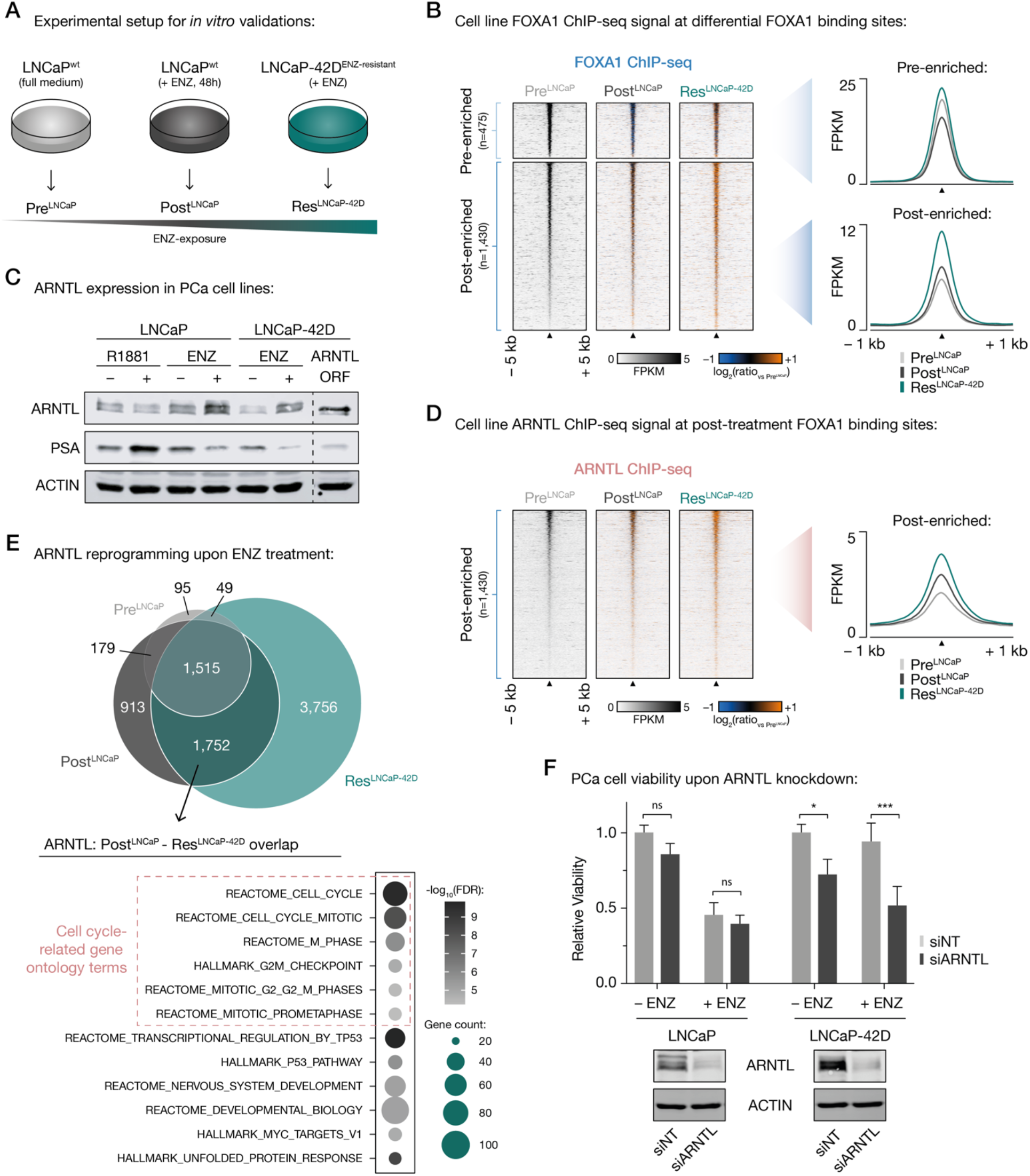
Treatment-induced dependency on ARNTL in ENZ-resistant PCa cells. (A) Experimental setup for *in vitro* validation experiments. (B) Tornado plots (left) and average density plots (right) visualizing FOXA1 ChIP-seq signal (in FPKM) at pre-enriched (top) and post-enriched (bottom) FOXA1 binding sites in untreated (Pre^LNCaP^), short-term ENZ-treated (Post^LNCaP^), and ENZ-resistant NE-like LNCaP cells (Res^LNCaP-42D^). Data are centered at differential FOXA1 peaks depicting a 5-kb (heatmaps) or 1-kb (density plots) window around the peak center. Heatmap color depicts the ChIP-seq signal compared to the untreated condition (Pre^LNCaP^), with blue indicating lower peak intensity and orange indicating higher peak intensity. Average of two biological replicates is represented. (C) Western blot showing ARNTL protein levels in LNCaP and LNCaP-42D cells following treatment with synthetic androgen (R1881) and/or ENZ (ENZ) for 48 h. ARNTL overexpression (in LNCaP-42D cells) as well as stainings for PSA and ACTIN are included as controls for antibody staining, hormonal treatment and protein loading, respectively. Images are representative of three independent experiments. (D) Tornado plots (left) and average density plot (right) visualizing ARNTL ChIP-seq signal (in FPKM) at post-enriched FOXA1 binding sites in untreated (Pre^LNCaP^), short-term ENZ-treated (Post^LNCaP^), and ENZ-resistant NE-like LNCaP cells (Res^LNCaP-42D^). Data are centered at post-treatment FOXA1 peaks depicting a 5-kb (heatmaps) or 1-kb (density plots) window around the peak center. Heatmap color depicts the ChIP-seq signal compared to the untreated condition (Pre^LNCaP^), with blue indicating lower peak intensity and orange indicating higher peak intensity. Average of two biological replicates is represented. (E) Venn diagram (top) indicating the overlap of ARNTL binding sites in all tested cell line conditions (Pre^LNCaP^, Post^LNCaP^, Res^LNCaP-42D^). For each condition, only peaks present in both replicates were included. Gene ontology terms for ARNTL- bound gene sets uniquely shared between Post^LNCaP^ and Res^LNCaP-42D^ conditions are presented below. Overlapping ARNTL binding sites (n=1,752) were coupled to their respective target genes using H3K27ac HiChIP data. Color indicates the gene set enrichment (FDR q-value) and size depicts the number of genes that overlap with the indicated gene sets. Cell cycle-related gene ontology terms are highlighted. (F) Bar chart (top) showing relative cell viability of LNCaP (left) and LNCaP-42D (right) cells upon transfection with non- targeting siRNA (siNT) or siARNTL, and exposure to ENZ. Cell viability assays were performed 7 days post-transfection. ENZ treatment is indicated and the average of three biological replicates is shown relative to the untreated (– ENZ) siNT condition per cell line. Western blots (bottom) indicate ARNTL protein levels in LNCaP (left) and LNCaP-42D (right) cells following siRNA-mediated silencing of ARNTL for 48 h. Transfection with siNT and staining for ACTIN are included as controls for siRNA treatment and protein loading, respectively. Images are representative of three independent experiments. ns, *P* > 0.05; *, *P* < 0.05; ***, *P* < 0.001 (two-way ANOVA followed by Tukey’s multiple comparisons test).

We performed FOXA1 ChIP-seq experiments in all three cell line conditions (**Supplementary Fig. S10A- S10D**; **Supplementary Table S6**), which revealed highly similar FOXA1 chromatin binding dynamics as observed in our clinical samples: While the pre-enriched FOXA1 sites identified *in vivo* showed less binding upon treatment, we observed that merely 48 h of ENZ exposure was sufficient to strongly induce binding at post-enriched sites, which was further increased in the long-term exposed, treatment-resistant LNCaP- 42D cell line (**Fig. 6B**; **Supplementary Fig. S10E**). Similarly, genome-wide correlation analyses indicated that short-term ENZ treatment in cell lines induced FOXA1 reprogramming to regions that are FOXA1- bound in treatment-resistant but not in treatment-naïve cells (**Supplementary Fig. S10F and S10G**).

Having shown that differential FOXA1 chromatin binding in tumors could be recapitulated *in vitro*, we next sought to further assess the role of ARNTL in these pre-clinical models. Therefore, we first confirmed that treatment with ENZ increased ARNTL protein levels in LNCaP and LNCaP-42D models (**Fig. 6C**), recapitulating the clinical observations (**Fig. 5C and 5D**). Since cistromic ARNTL profiling has to date not been reported in PCa models, we generated ARNTL ChIP-seq data (**Supplementary Fig. S11A-S11D**) to validate its binding at post-treatment FOXA1 sites. Interestingly, while we already observed ARNTL binding to these regulatory regions in the pre-treatment setting, this was strongly enhanced upon ENZ exposure (**Fig. 6D**; **Supplementary Fig. S11E**). To identify functional differences in ARNTL cistromes induced upon treatment, we overlapped the ARNTL peaks identified in all tested cell line conditions, which revealed a massive cistromic reprogramming upon ENZ treatment (**Fig. 6E**; **Supplementary Fig. S11F and S11G**). Notably, ∼70% of ENZ-gained ARNTL peaks (n=1,752) in LNCaP cells were captured by the ARNTL cistrome in treatment-resistant cells. Subsequent pathway over-representation analyses of genes coupled to these Post^LNCaP^-Res^LNCaP-42D^-shared ARNTL binding sites revealed a treatment-induced enrichment for gene sets implicated in cell cycle progression and cell division, further supporting a possible functional involvement of ARNTL in sustaining tumor cell proliferation when AR is blocked by ENZ (**Fig. 6E**). To challenge this hypothesis, we assessed whether ARNTL-knockdown affects the viability of hormone-sensitive and in particular of long-term ENZ-exposed cell lines. While ARNTL-targeting had minimal effect on LNCaP cell proliferation (with or without ENZ), ARNTL knockdown significantly suppressed cell growth of ENZ-resistant LNCaP-42D cells in the absence (*P* = 0.031, two-way ANOVA) and even more so in the presence of ENZ (*P* = 7 x 10^-4^, two-way ANOVA), indicating that targeting ARNTL also partially restores ENZ-sensitivity in this treatment-resistant cell line model (**Fig. 6F**).

Overall, these data confirm the ENZ-induced FOXA1 reprogramming as observed in PCa patients upon neoadjuvant antiandrogen therapy, and revealed an acquired dependency on circadian rhythm regulator ARNTL to drive tumor cell growth – positioning ARNTL as a highly promising new drug target in combination with ENZ for the treatment of high-risk PCa.

## Discussion

In medicine, the evolutionary selection pressure as imposed by drug treatment has been a well-known clinical challenge, ever since the first antibiotics were discovered in the early 20th century. Also in oncology, clear escape mechanisms for both targeted therapeutics and systemic treatments are known for many years, involving *ESR1* mutations in metastatic breast cancer (44), *EGFR* mutations in lung cancer (45), *KRAS* mutations in metastatic colorectal cancer (46), but also somatic amplification of the *AR* locus and/or an upstream *AR* enhancer in castration-resistant PCa (47, 48). Apart from genetic alterations, also epigenetic rewiring (7, 41) or transdifferentiation are reported as mechanisms of resistance, including treatment-emergent neuroendocrine (NE) prostate cancers that occur as an adaptive response under the pressure of prolonged AR-targeted therapy (49, 50).

Our unique clinical trial design with paired pre- and post-treatment biopsies of high-risk primary PCa treated with ENZ monotherapy, allowed us to unravel global ENZ-induced alterations in gene regulation. We report that large-scale treatment-induced dedifferentiation in PCa may be a gradual process, of which the early signs are identified on transcriptomic level within the first months of treatment onset. While complete adenocarcinoma-NE transdifferentiation was not observed in any of our samples, cellular plasticity characterized by transcriptomic features of NE disease may not only be present prior to treatment (31), but also become enriched upon short-term endocrine treatment exposure.

In PCa development (5, 51) and progression (7), AR has been reported to expose substantial plasticity in its enhancer repertoire, and we now illustrate this is also the case in primary disease upon short-term treatment. Besides AR, FOXA1 is considered a master transcription factor and critical prostate lineage specific regulator acting in PCa, that upon overexpression during tumorigenesis gives rise to a tumor-specific AR cistrome. Also in NEPC, FOXA1 cistromes are reprogrammed (52), which indicates a direct AR-independent role of FOXA1 in PCa progression. Our study confirms these observations and shows that, while co-occupied by AR, the pre- and post-ENZ enriched FOXA1 sites appeared indifferent to AR signaling.

The functional implications of the pre-treatment FOXA1 sites remain unclear, as those regions were inactive, both in primary tissues as well as in reporter assays. A subset of these *cis*-regulatory elements demarcates developmental epigenomic programs, that we previously reported as being occupied by FOXA1 from prostate development to tumorigenesis and metastatic progression (7), whereas others may be relevant for different physiological processes.

The treatment-induced cistromic repositioning of FOXA1 initiated a thus far unknown transcriptional rewiring, in which ARNTL, a classical circadian rhythm regulator and dimerization partner of CLOCK, compensates for AR inhibition and becomes essential to rescue cellular proliferation signals. Recently, it has been reported that CRY1 – a transcriptional coregulator of ARNTL – is AR-regulated in PCa, and modulates DNA repair processes in a circadian manner (53). The current data illustrate that circadian rhythm may have a potential impact on drug response, as most clock components are regulated on transcriptional level, in time. Our data now show that AR blockade forces tumor cells to adapt epigenetically, upon which these cells – over time – become dependent on ARNTL as a transcriptional regulator of proliferation processes. This acquired cellular vulnerability appears to be dependent on whether or not AR activity is inhibited and cells have had time to achieve full epigenetic reprogramming, explaining the limited effect of ARNTL knockdown in hormone-sensitive PCa cells, as compared to the long-term ENZ-exposed treatment-resistant model.

With the identification of ARNTL as a rescue mechanism for tumor cells to evade AR blockade, the next question presents whether ARNTL could serve as a novel therapeutic target, which should be further pursued in future drug development and clinical research. Being critically relevant for circadian rhythm regulation, it would be imperative to balance ARNTL targeting in relation to any adverse side-effects. Additionally, we demonstrate that the surprisingly dynamic enhancer repertoire of FOXA1 is not only critical in prostate tumorigenesis (5) and neuroendocrine differentiation (52), but also appears crucial in evading AR therapy-induced growth inhibition, further supporting the rationale to intensify efforts in targeting this highly tissue-selective, yet critical transcriptional regulator, directly or indirectly (54).

## Methods

### Study design

Primary PCa tissues before and after enzalutamide (ENZ) treatment were acquired as part of the phase 2, prospective, single-arm DARANA study (ClinicalTrials.gov #NCT03297385) at the Netherlands Cancer Institute Antoni van Leeuwenhoek hospital. To allow sample size calculation, we performed a survey into the surgical margins of 1492 in-house prostatectomy specimens (Gleason ≥7), not treated with antihormonal therapy prior to surgery, which revealed 34% not-radical resections. Earlier randomized studies on neoadjuvant androgen ablation showed reductions in positive surgical margin rate of at least 50% (55–57). To detect a reduction of positive surgical margins from 34% to 17% with a power of 80% and an alpha set at 0.05, 55 patients needed to be included. Inclusion criteria were over 18 years of age, Gleason ≥ 7 PCa and planned for prostatectomy. Prior to treatment a multi-parametric MRI scan was made to identify tumors in the prostate (cT-stage) and pelvic lymph node metastasis (cN-stage). Patients were treated with ENZ, once daily 160 mg P.O. without androgen deprivation therapy, for three months prior to RALP and a pelvic lymph node dissection. 55 patients completed therapy without dose adjustments, while one patient (DAR37) discontinued ENZ three weeks prematurely. The resection specimen was assessed for prostate tumor stage (ypT-stage) and the pelvic lymph nodes (ypN-stage). Primary clinical outcome measure was positive margins rate and secondary endpoints were differences in pre- and post-treatment T (tumor) and N (lymph node) stage and median time to biochemical recurrence, defined as two consecutive rises of serum PSA with a minimal level of ≥ 0.2 ng/mL. The trial was approved by the institutional review board of the Netherlands Cancer Institute, informed consent was signed by all participants enrolled in the study, and all research was carried out in accordance with relevant guidelines and regulations.

### Pre- and post-treatment sampling

Prior to ENZ intervention, 4 pre-operative MRI-guided 18-gauche core needle tumor biopsies were taken per patient. Directly after prostatectomy, 8 additional tumor-targeted core needle biopsies (4x 14 gauche, 4x 5 mm) were taken from prostatectomy specimens *ex vivo*, using previous MRI information and palpation. Biopsy and prostatectomy specimens were fresh frozen (FF) or formalin-fixed paraffin-embedded (FFPE) for ChIP-seq and CNV-seq, or RNA-seq and immunohistochemistry analyses, respectively. Prior to ChIP-seq experiments, FF material was cut in 30 µm sections, while FFPE material was cut in 10 µm sections prior to RNA extraction. Tissue sections were examined pathologically for tumor cell content and only samples with a tumor cell percentage of ≥50% were used for further downstream analyses.

### ChIP-seq

Chromatin immunoprecipitations on PCa tissue specimens and cell line models were performed as previously described (58). In brief, cryosectioned tissue samples were double-crosslinked in solution A (50 mM HEPES-KOH, 100 mM NaCl, 1 mM EDTA, 0.5 mM EGTA), first supplemented with 2 mM disuccinimidyl glutarate (DSG; CovaChem) for 25 min at room temperature. Then, 1% formaldehyde (Merck) was added for 20 min and subsequently quenched with a surplus of 2.5 M glycine. Cell lines were crosslinked using single-agent fixation. Therefore, 1% formaldehyde was added to the cell culture medium and incubated at room temperature for 10 min, followed by glycine-quenching as described above. Tissue and cell line samples were lysed as described (59) and sonicated for at least 10 cycles (30 sec on; 30 sec off) using a PicoBioruptor (Diagenode). For each ChIP, 5 µg of antibody were conjugated to 50 µL magnetic protein A or G beads (10008D or 10009D, Thermo Fisher Scientific). The following antibodies were used: AR (06-680, Merck Millipore), FOXA1 (ab5089, Abcam), H3K27ac (39133, Active Motif), and ARNTL (ab93806, Abcam).

Immunoprecipitated DNA was processed for library preparation using a KAPA library preparation kit (KK8234, Roche) and generated libraries were sequenced on the Illumina HiSeq2500 platform using the single end protocol with a read length of 65-bp, and aligned to the human reference genome hg19 using Burrows-Wheeler Aligner (v0.5.10) (60). Reads were filtered based on mapping quality (MAPQ ≥ 20) and duplicate reads were removed. Peak calling over input controls (per tissue sample or cell line) was performed using MACS2 (v2.1.1) and Dfilter (v1.6) for tissues, and MACS2 (v2.1.2) for cell lines (61, 62). For tissue samples, only the peaks shared by both peak callers were used for downstream analyses. DeepTools (v2.5.3) was used to calculated read counts in peaks (FRiP) (63). Read counts and the number of aligned reads, as well as normalized strand coefficient (NSC) and relative strand correlation (RSC), which were calculated using phantompeaktools (v1.10.1) (64), are shown in **Supplementary Table S2** for tissue ChIP-seq data and **Supplementary Table S6** for cell line ChIP-seq data. Tissue ChIP-seq samples that passed the following quality control measures were included in the final analyses; tumor cell percentage ≥ 50%, ChIP-qPCR enrichment, and more than 100 peaks called (**Supplementary Fig. S1B**).

For visualization of cell line ChIP-seq data, an average enrichment signal was generated by merging mapped reads of replicate samples using SAMtools (v1.10-3) (65).

Genome browser snapshots, tornado and average density plots were generated using EaSeq (v1.101) (66). Genomic distribution and motif enrichment analyses were performed using the CEAS and the SeqPos motif tools on Galaxy Cistrome (67), respectively. Cistrome Toolkit was used to probe which TFs and chromatin regulators have a significant binding overlap with the differential FOXA1 peak sets (39). For this, genomic coordinates of high-confidence binding sites (FC ≥ 1.2) were converted between assemblies (from hg19 to hg38), using the UCSC genome browser liftOver tool (68). The DiffBind R package (v2.10) was used to generate correlation heatmaps and PCA plots based on occupancy, to perform differential binding analyses using a false discovery rate (FDR) < 0.05, and to generate consensus peaklists (69).

ChIP-seq signal of various datasets (FOXA1, AR and H3K27ac from this study; AR, H3K27ac and H3K27me3 from a previously reported study (31)) at differential and consensus FOXA1 sites was investigated by counting mapped reads in FOXA1 peak regions using bedtools multicov (v2.27.1) (70). Readcounts were subsequently z-transformed and visualized using the aheatmap function from the R package NMF (v0.21.0) (71) with a color scheme from RColorBrewer (v1.1-2; https://CRAN.R-project.org/package=RColorBrewer). To determine significance in binding site occupancy differences between pre- and post-treatment FOXA1 sites, median z-transformed readcounts were calculated per sample and compared using a Mann-Whitney U-test. These median readcounts per sample were also used to assess the correlation between ChIP-seq signals of AR, FOXA1 and H3K27ac at pre-enriched, post-enriched and consensus FOXA1 binding sites.

Bedtools intersect (v2.27.1) (70) was used to determine overlap of differential FOXA1 binding sites and inactive, constitutively active and inducible AR-binding sites.

To assign FOXA1 and ARNTL binding regions to potential target genes, we overlapped differential FOXA1 binding sites with H3K27ac HiChIP data (35) using bedtools intersect. To assess whether or not genes coupled to FOXA1 binding sites were considered to be essential for the VCAP prostate cancer cell line, we used the DepMap (Broad 2020) 20Q1 Public gene effect dataset (36). Gene set overlaps between genes linked to ChIP-seq binding sites and the Molecular Signatures Database (v7.4) were computed using Gene Set Enrichment Analysis (GSEA) (72) with an FDR q-value cutoff ≤ 0.05.

### RNA-seq

Prior to RNA isolation, FFPE material was pathologically assessed. The expert pathologist scored tumor cell percentage and indicated most tumor-dense regions for isolation on a hematoxylin and eosin (H&E) slide. RNA and DNA from FFPE material were simultaneously isolated from 3-10 sections (depending on tumor size) of 10 µm using the AllPrep DNA/RNA FFPE isolation kit (80234, Qiagen) and the QIAcube according to the manufacturer’s instructions. cDNA was synthesized from 250 ng RNA using SuperScript III Reverse Transcriptase (Invitrogen) with random hexamer primers.

For RNA-seq, strand-specific libraries were generated with the TruSeq RNA Exome kit (Illumina) and sequenced on the Illumina HiSeq2500 platform using the single end protocol with a read length of 65-bp.

Sequencing data was aligned to the human reference genome hg38 using TopHat (v2.1.0 using bowtie 1.1.0) (73) and the number of reads per gene was measured with HTSeq count (v0.5.3) (74).

For QC purposes, total readcounts per sample were determined and hierarchical clustering based on the Euclidean distance was applied. Samples with a readcount ≥ 2 standard deviations below the mean of all sample readcounts were removed, as well as samples that clustered in a separate branch.

Global gene expression differences between pre- and post-treatment samples passing QC were determined using DESeq2 (v1.22.2) (75). Significance of expression level differences between pre- and post-treatment samples was determined using a paired t-test.

Gene set enrichment was performed using pre-ranked GSEA (72) based on the Wald statistic provided by DESeq2. For visualization purposes, the data were Z-transformed per gene. Heatmaps of gene expression values were created using the aheatmap function from the R package NMF (v0.21.0) (71) with a color scheme from RColorBrewer (v1.1-2; https://CRAN.R-project.org/package=RColorBrewer).

To assign samples to previously described PCa subtypes (31), the z-transformed expression levels of the top ∼100 most differentially expressed genes (n=285) in each of the three clusters were investigated. Using these values, samples were clustered based on their Pearson correlation. The resulting tree was divided into 3 clusters, corresponding to the previously published PCa subtypes. Potential transitioning of samples from one cluster to another after treatment was visualized using a riverplot (v0.6; https://CRAN.R-project.org/package=riverplot).

To calculate fold changes of neuroendocrine scores upon treatment, expression of 70 neuroendocrine signature genes were obtained from castration-resistant neuroendocrine and prostate adenocarcinoma samples as published previously (34). The expression of 5 of the 70 neuroendocrine signature genes were not included in the analysis (KIAA0408, SOGA3, LRRC16B, ST8SIA3, SVOP) because the genes are not expressed in these samples. Expression fold changes between paired pre- and post-treatment samples were calculated (n=39) and concordance in gene expression differences (fold change sign) were measured using Pearson correlation.

### CNV-seq

Low-coverage whole-genome samples (ChIP-seq inputs), sequenced single-end 65-bp on a HiSeq 2500 system were aligned to hg19 with Burrows-Wheeler Aligner backtrack algorithm (v0.5.10) (60). The mappability per 20-kb on the genome, for a samples’ reads, phred quality 37 and higher, was rated against a similarly obtained mappability for all known and tiled 65-bp subsections of hg19. Sample counts were corrected per bin for local GC effects using a non-linear loess fit of mappabilities over 0.8 on autosomes. Reference values were scaled according to the slope of a linear fit, forced to intercept at the origin, of reference mappabilities after GC correction. Ratios of corrected sample counts and reference values left out bins with mappability below 0.2 or overlapping ENCODE blacklisted regions (76).

Copy number log ratios were smoothed and segmented using the R package DNACopy (v1.50.1; https://bioconductor.org/packages/release/bioc/html/DNAcopy.html) with the parameters set to alpha=0.00000000001, undo.SD=2, and undo.splits=“sdundo”. Bedtools intersect (v2.27.1) (70) was used to determine overlap between copy number segments and differential FOXA1 binding sites. These data were subsequently visualized using the aheatmap function from the R package NMF (v0.21.0) (71) with a color scheme from RColorBrewer (v1.1-2; https://CRAN.R-project.org/package=RColorBrewer).

To correlate FOXA1 ChIP-seq signal with copy number status at differential FOXA1 sites, we employed the z-transformed FOXA1 ChIP-seq readcounts as described in the ChIP-seq section. The difference in transformed ChIP-seq readcounts and the difference in normalized segmented copy number data between matched post-treatment and pre-treatment samples was calculated for every patient. Subsequently, the Pearson correlation between these two sets of differences was calculated.

### Immunohistochemistry

For immunohistochemistry (IHC) analysis, we matched our ENZ-treated patient cohort (n=51) in a 1:2 ratio to untreated control patients (not receiving ENZ prior to prostatectomy; n=110) based on clinicopathological parameters (initial PSA, Gleason score, TNM stage, age) using the R package MatchIt (v.4.1.0) (77).

Tissue microarrays (TMAs) were prepared containing 3 cores per FFPE tumor sample. Tumor-dense areas in FFPE megablocks were marked by an expert pathologist on a H&E slide. Cores were drilled in a receptor block using the TMA grandmaster (3D Histech/Sysmex). Next, cores were taken from the donor block and placed in the receptor block using the manual tissue arrayer (4508-DM, Beecher instruments). The filled receptor block was placed in a 70°C stove for 9 minutes and cooled overnight at RT. Immunohistochemistry was applied to TMA slides using a BenchMark Ultra autostainer (Ventana Medical Systems). In brief, paraffin sections were cut at 3 µm, heated at 75°C for 28 minutes and deparaffinized in the instrument with EZ prep solution (Ventana Medical Systems). Heat-induced antigen retrieval was carried out using Cell Conditioning 1 (CC1, Ventana Medical Systems) for 64 minutes at 95°C.

For ARNTL IHC, TMAs were stained with an anti-ARNTL antibody (ab230822, Abcam) for 60 minutes at 36°C using a 1:1000 antibody dilution. Bound antibody was detected using the OptiView DAB Detection Kit (Ventana Medical Systems). Slides were counterstained with hematoxylin and bluing reagent (Ventana Medical Systems).

ARNTL staining intensity (weak, moderate, strong) in tumor cells was scored by an expert pathologist. Tissues scored for at least two cores were analyzed and used for statistical analysis.

### Cell lines and cell culture

LNCaP human PCa cell line was purchased from the American Type Culture Collection (ATCC). Enzalutamide-resistant LNCaP-42D cells were described previously (43). LNCaP cells were maintained in RPMI-1640 medium (Gibco, Thermo Fisher Scientific) supplemented with 10% FBS (Sigma-Aldrich). LNCaP-42D cells were maintained in RPMI-1640 medium containing 10% FBS and 10 µM ENZ. Cell lines were subjected to regular mycoplasma testing and all cell lines underwent authentication by short tandem repeat profiling (Eurofins Genomics). For hormone stimulation with synthetic androgen, cells were treated with 10 nM R1881 (PerkinElmer) for 48 h. For *in vitro* AR blockade, cells were treated with 10 µM ENZ (MedChemExpress) and harvested at the indicated time points.

### Transient cell line transfections

Transient transfections of cell lines were performed according to the manufacturer’s instructions using Lipofectamine 2000 (Invitrogen) or Lipofectamine RNAiMAX (Invitrogen) for overexpression or siRNA knockdown experiments, respectively. ARNTL containing expression plasmid was obtained from the CCSB-Broad Lentiviral Expression Library. siRNA oligos targeting ARNTL (M-010261-00-0005), and the non-targeting control (D-001206-14) were purchased from Dharmacon.

### Western blotting

Total proteins were extracted from cells using Laemmli lysis buffer, supplemented with a complete protease inhibitor cocktail (Roche). Per sample, 40 µg of protein was resolved by SDS-PAGE (10%) and transferred on nitrocellulose membranes (Santa Cruz Biotechnology). The following antibodies were used for Western blot stainings: ARNTL (ab93806, Abcam), PSA (5365, Cell Signaling Technology), and ACTIN (MAB1501R, Merck Millipore). Blots were incubated overnight at 4°C with designated primary antibodies at 1:1000 (ARNTL, PSA) or 1:5000 (ACTIN) dilution, and visualized using the Odyssey system (Li-Cor Biosciences).

### Cell viability assay

For cell viability assays, LNCaP or LNCaP-42D cells were seeded at 2 x 10^3^ cells per well in 96-well plates (Greiner) ± 10 µM ENZ, and reverse transfected with 50 nM siRNA (Dharmacon) using Lipofectamine RNAiMAX (Invitrogen). Cell viability was assessed 7 days post-transfection using the CellTiter-Glo Luminescent Cell Viability Assay kit (Promega), as per the manufacturer’s instructions.

### Statistical analysis

For differential binding and differential gene expression analyses (pre- vs. post-ENZ), an FDR cutoff < 0.05 (*P* < 0.01) and *Padj* < 0.01 was used, respectively. A Mann-Whitney U-test was used to determine differences in region readcounts (adjusted for multiple testing using FDR) and differences in gene expression levels before and after ENZ treatment. For peak set and gene set overlaps, Fisher’s exact or Welch Two Sample t-tests were applied. A Fisher’s exact test was used to determine differences in IHC staining intensity. Differences in cell viability were tested using a two-way ANOVA followed by Tukey’s multiple comparisons test (GraphPad Prism 9). Corresponding bar chart shows the mean with error bars representing the SD of three independent experiments. All boxplots indicate the median (center line), upper-(75) and lower - (25) quartile range (box limits) and 1.5 x interquartile range (whiskers). Significance is indicated as follows: ns, *P* > 0.05; *, *P* < 0.05; **, *P* < 0.01; ***, *P* < 0.001; ****, *P* < 0.0001. Further details of statistical tests are provided in the figure legends.

## Supporting information

Supplementary Table S1 - Clinical outcome

Supplementary Table S2 - ChIP-seq quality control metrics

Supplementary Table S3 - Differential FOXA1 binding sites

Supplementary Table S4 - Intrinsic enhancer activity of tumor-specific ARBS

Supplementary Table S5 - List of genes coupled to differential FOXA1 sites based on H3K27ac HiChIP enhancer-promoter coupling

Supplementary Table S6 - Cell line ChIP-seq quality control metrics

## Data Availability

All data produced in the present study are available upon reasonable request to the authors.

## Acknowledgements

The authors would like to thank the NKI-AVL Core Facility Molecular Pathology & Biobanking (CFMPB) for technical assistance, the NKI Genomics Core Facility (GCF) for next generation sequencing and bioinformatics support, and the NKI Research High Performance Computing facility (RHPC) for computational infrastructure. We also thank all Zwart/Bergman lab members for fruitful discussions and technical advice. Finally, we would like to thank all patients and clinical staff who were involved in the DARANA trial.

