## Supplementary Data and Figures for "Drug-induced epigenomic plasticity reprograms circadian rhythm regulation to drive prostate cancer towards androgen-independence"

#### Clinical trial design and primary clinical outcomes

We conducted a single-arm, open-label Phase II clinical trial: the DARANA study (Dynamics of Androgen Receptor Genomics and Transcriptomics After Neoadjuvant Androgen Ablation; NCT03297385). In this study, 56 men with primary high-risk (Gleason score ≥ 7) PCa were enrolled. Mean age at inclusion was 67 years, baseline serum PSA 12.8 ng/mL and the majority of patients had an ISUP2 (28%) or ISUP4 (36%) PCa (**Fig. 1A**; **Table 1**). Prior to ENZ therapy, magnetic resonance imaging (MRI)-guided core needle tumor biopsies were taken. Subsequently, patients received neoadjuvant ENZ treatment (160 mg/day) for three months without androgen deprivation therapy. 55 patients completed therapy without dose adjustments, while one patient (DAR37) discontinued ENZ three weeks prematurely. All patients underwent a robotic-assisted laparoscopic prostatectomy (RALP). Based on baseline MRI information and palpation, additional tumor-targeted core needle biopsies were taken *ex vivo*. 30% of prostatectomy specimens had positive surgical margins, which was comparable to 1492 non-treated historical controls with Gleason ≥7 (34%; see methods) and analogous to a previous neoadjuvant study on ENZ alone versus ENZ in combination with the 5$\alpha$ -reductase inhibitor dutasteride and androgen deprivation therapy in high-risk patients for 6 months, describing 26.1% positive margins in the triple-therapy arm (1). No differences in pre and post treatment T-stages were observed. A higher incidence of ypN1 lymph nodes than cN1 lymph nodes was possibly due to differences in accuracy of radiographic pre-treatment assessments versus post-treatment histological assessments. After a mean follow-up of 37 months, the mean time to biochemical recurrence was 12 months (**Supplementary Fig. S1A**; **Supplementary Table S1**).

### Supplementary Figures

#### Supplementary Figure S1

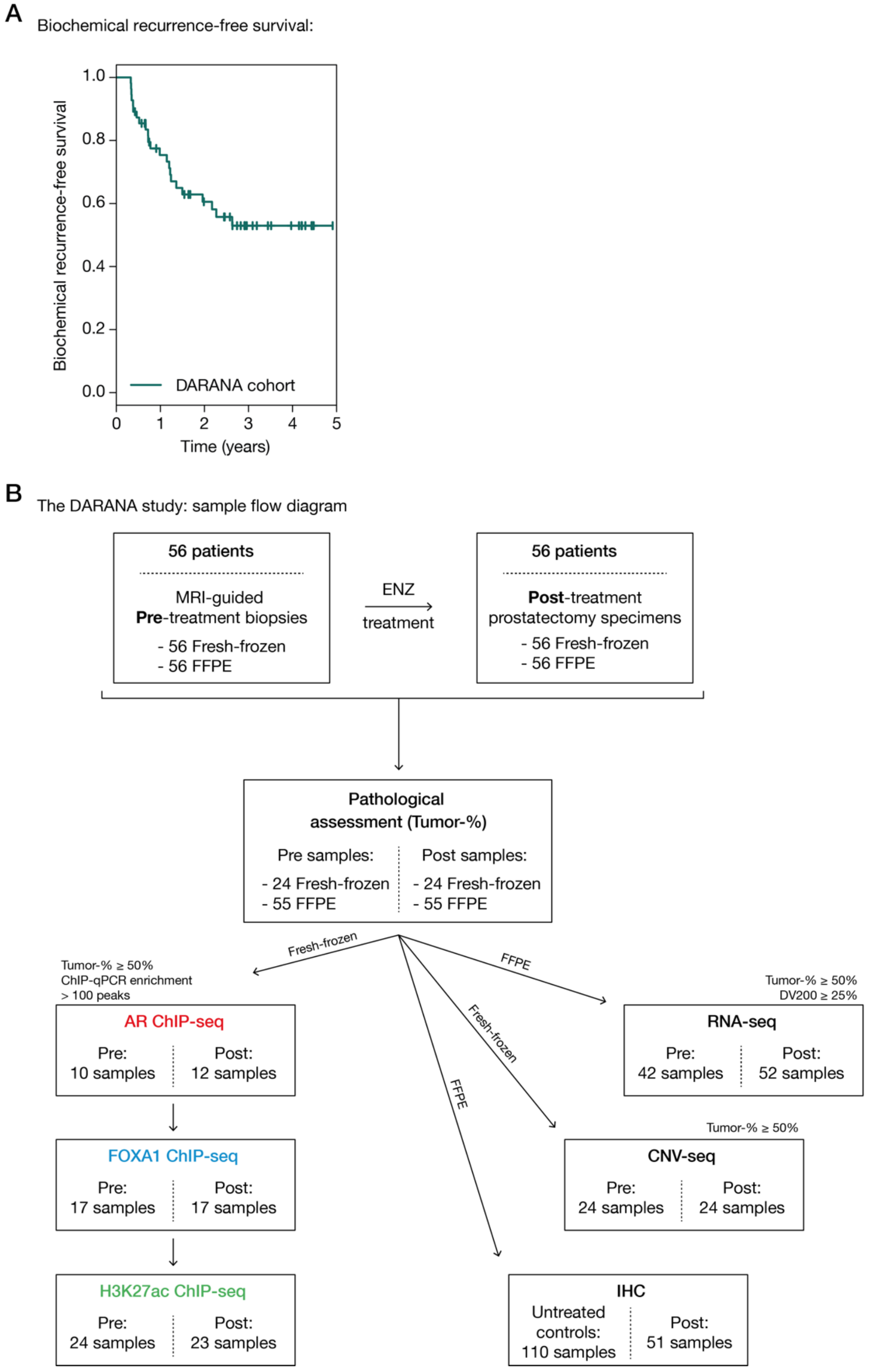


**Figure S1: The DARANA study.**

(A) Kaplan-Meier curve showing the biochemical recurrence (BCR)-free survival of the DARANA cohort, treated with 3 months of neoadjuvant ENZ prior to prostatectomy. BCR was defined as a rise in Prostate-specific antigen (PSA) serum levels of $\geq 0.2$ ng/mL.

(B) Sample flow diagram indicating the quality control measures applied to each sample, and the number of samples passing the respective cut-offs per omics data stream.

#### Supplementary Figure S2

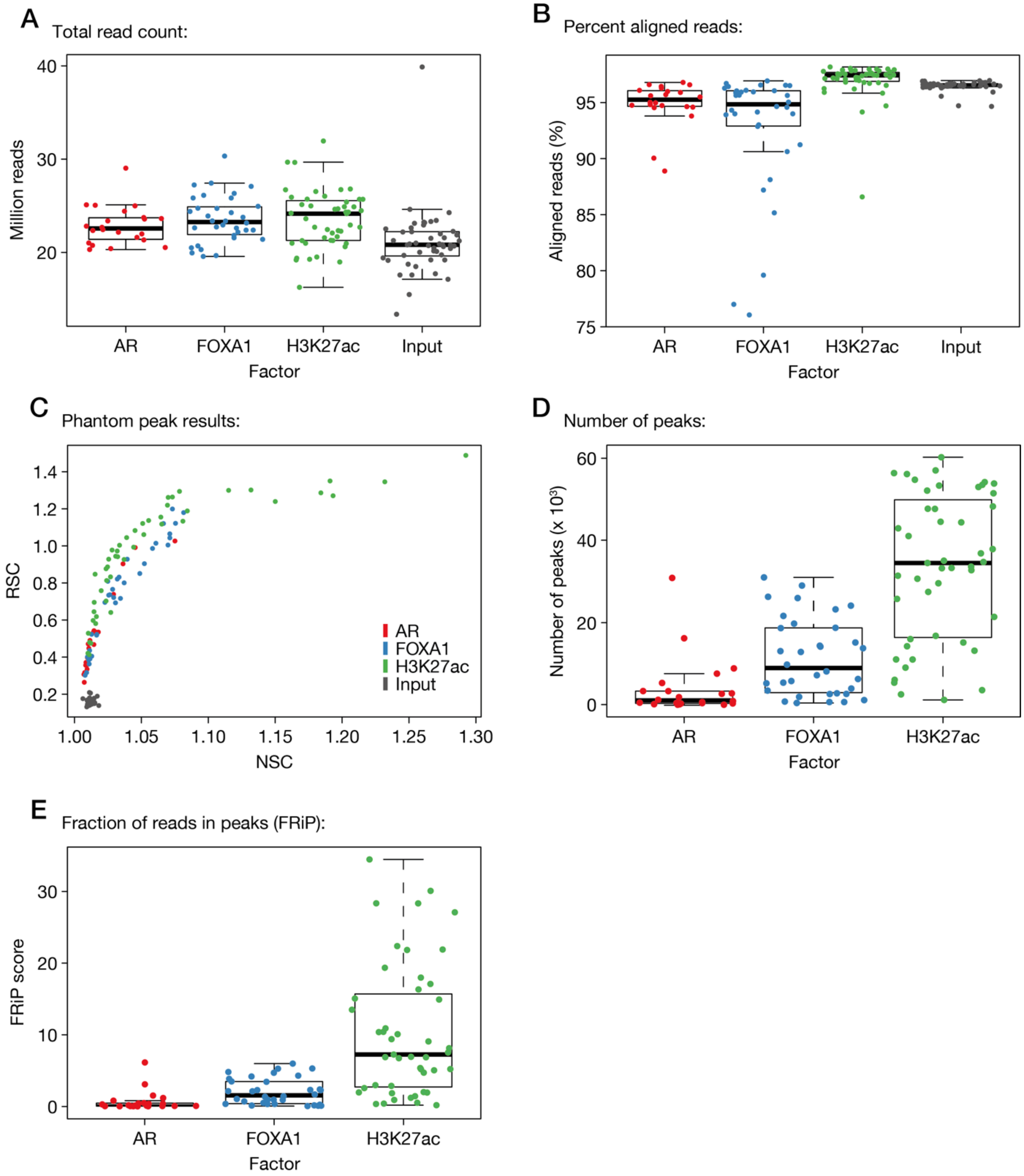


**Figure S2: Tissue ChIP-seq quality metrics.**

(A) Boxplots showing the median total number of reads for AR (red), FOXA1 (blue) and H3K27ac (green) ChIP-seq, as well as ChIP-seq input (gray) samples.

(B) Boxplots indicating the percentage of aligned reads per ChIP factor.

(C) Scatter plot of phantom peak results, showing the normalized strand cross-correlation coefficient (NSC) and relative strand cross-correlation coefficient (RSC) scores for each ChIP-seq and input sample.

(D) Boxplot showing the number of peaks per ChIP-seq sample. Peak calling was performed over matched input control samples.

(E) Boxplots indicating the fraction of reads in peaks (FRiP) score per sample.

#### Supplementary Figure S3

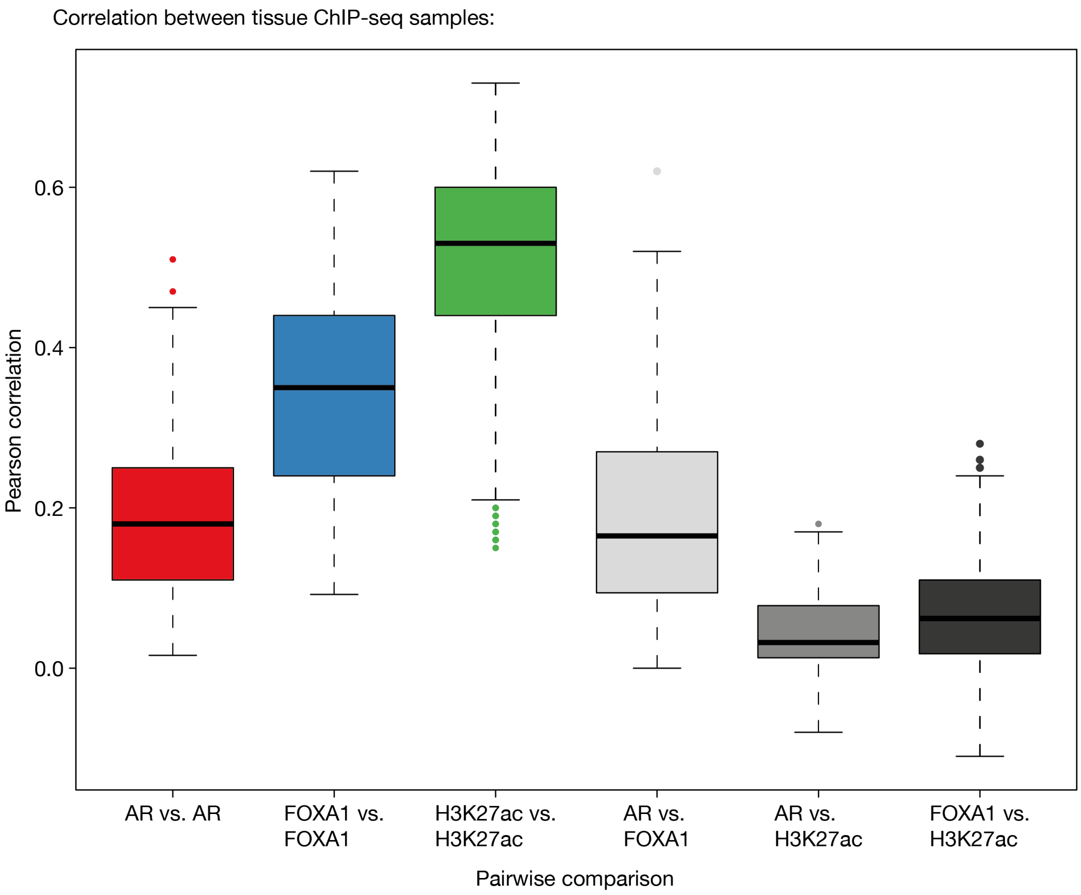


**Figure S3: Correlation between tissue ChIP-seq samples.**

Boxplots indicating the Pearson correlation of ChIP-seq samples based on peak occupancy. Both, pairwise comparisons within the same ChIP-seq dataset (AR vs. AR, FOXA1 vs. FOXA1, H3K27ac vs. H3K27ac), and between different datasets (AR vs. FOXA1, AR vs. H3K27ac, FOXA1 vs. H3K27ac) are shown. The corresponding correlation matrix is shown in **Fig. 2B**.

#### Supplementary Figure S4

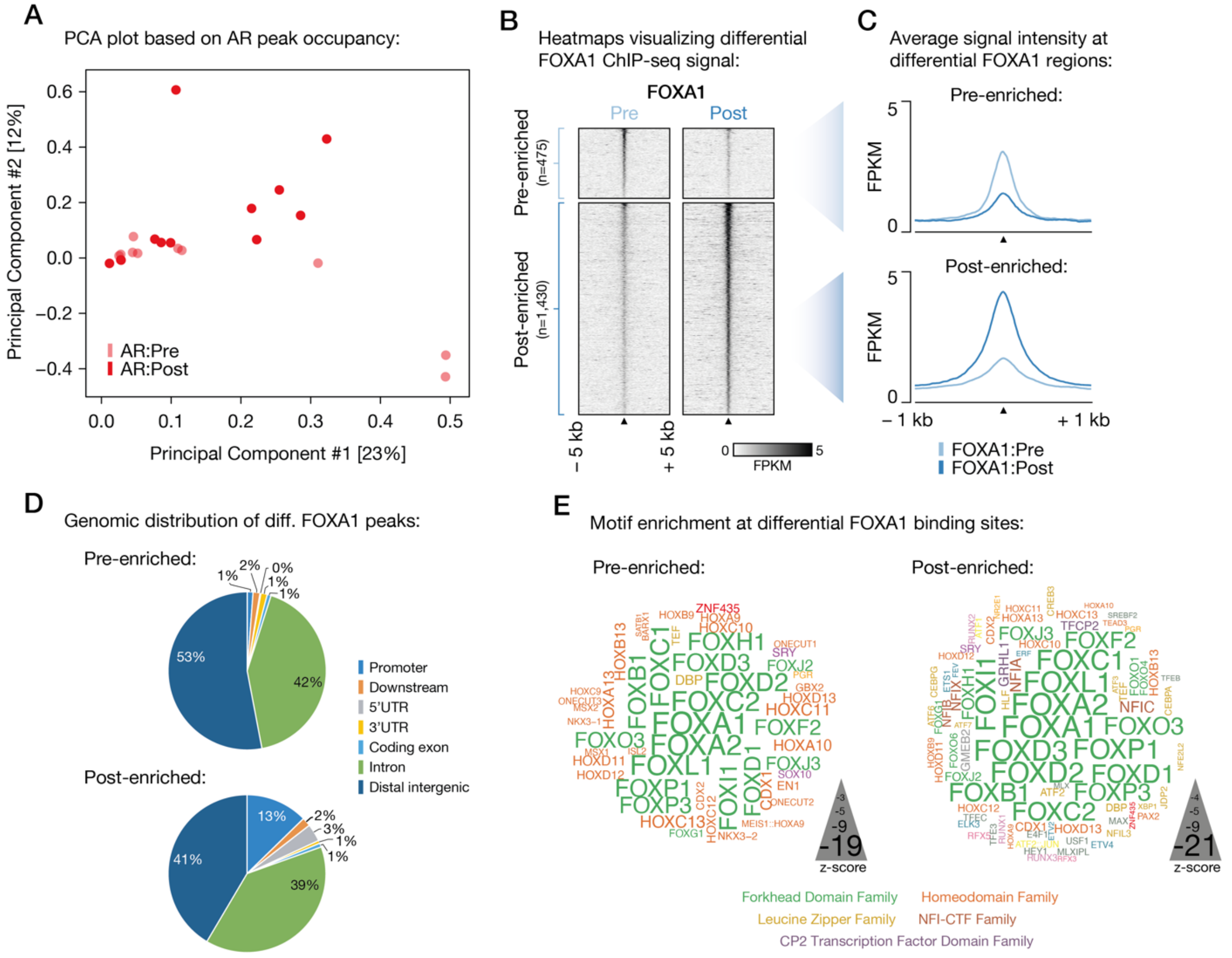


**Figure S4: Differential FOXA1 chromatin binding upon ENZ treatment.**

(A) Principle component analysis (PCA) plot based on peak occupancy of AR ChIP-seq data. Color indicates the treatment status of pre- (light red) and post- (dark red) ENZ AR samples.

(B) Representative tornado plots visualizing FOXA1 ChIP-seq signal (in fragments per kilobase per million reads mapped (FPKM)) at pre-enriched and post-enriched FOXA1 binding sites before (pre) and after (post) ENZ treatment in one patient. Data are centered at FOXA1 peaks depicting a 5-kb window around the peak center.

(C) Quantification of the average signal intensity at pre-enriched (top) and post-enriched (bottom) FOXA1 binding sites before (light blue) and after (dark blue) ENZ treatment shown in (B). Data are centered at FOXA1 peaks depicting a 2.5-kb window around the peak center.

(D) Pie charts showing the genomic distribution of pre-enriched (top) and post-enriched (bottom) FOXA1 binding sites.

(E) Word clouds show motif enrichment at pre-enriched (left) and post-enriched (right) FOXA1 sites. The font size represents the z-score and colors correspond to transcription factor families.

#### Supplementary Figure S5

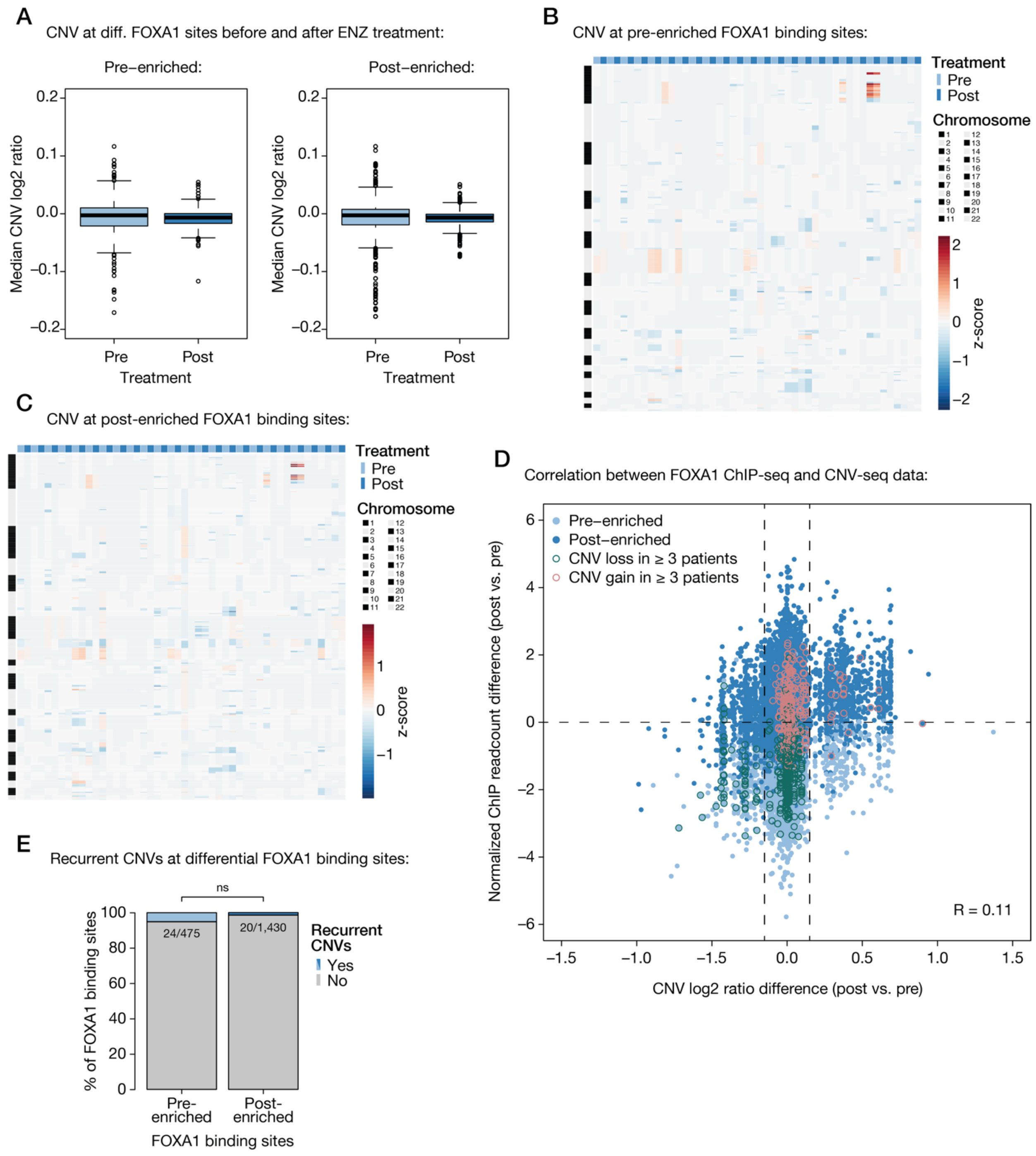


**Figure S5: Copy number variation at differential FOXA1 sites.**

(A) Boxplot showing the median copy number ratio at pre-enriched (left) and post-enriched (right) FOXA1 binding sites over all pre-treatment and post-treatment samples.

(B-C) Heatmap depicting segmented CNV data at pre-enriched (B) and post-enriched (C) FOXA1 binding sites in pre- and post-treatment samples (columns). Differential FOXA1 sites are ordered per chromosome (rows) and heatmap color indicates copy number gains and losses (z-score).

(D) Scatter plot showing the correlation between FOXA1 ChIP-seq data and CNV-seq data for paired pre-treatment and post-treatment samples (n=15). Plotted is the normalized ChIP readcount difference (post vs. pre ENZ) against the CNV ratio difference (post vs. pre ENZ) at all differential FOXA1 binding sites (n=1,905) per patient. CNV cut-offs of ± 0.1 (dotted vertical lines) and mean Pearson correlation ($R$ = 0.11; range: -0.02 – 0.26) are indicated. Differential FOXA1 binding sites (dots) are colored based on their enrichment (pre-enriched vs. post-enriched) and recurrent CNV events found in ≥ 3 patients are highlighted.

(E) Stacked bar plot indicating the fraction of differential FOXA1 sites that show recurrent CNVs shown in (D) with CNV ratio > 0.1 for post-enriched and < 0.1 for pre-enriched FOXA1 sites in 3 or more patients (pre: n=24/475, post: n=20/1,430).

#### Supplementary Figure S6

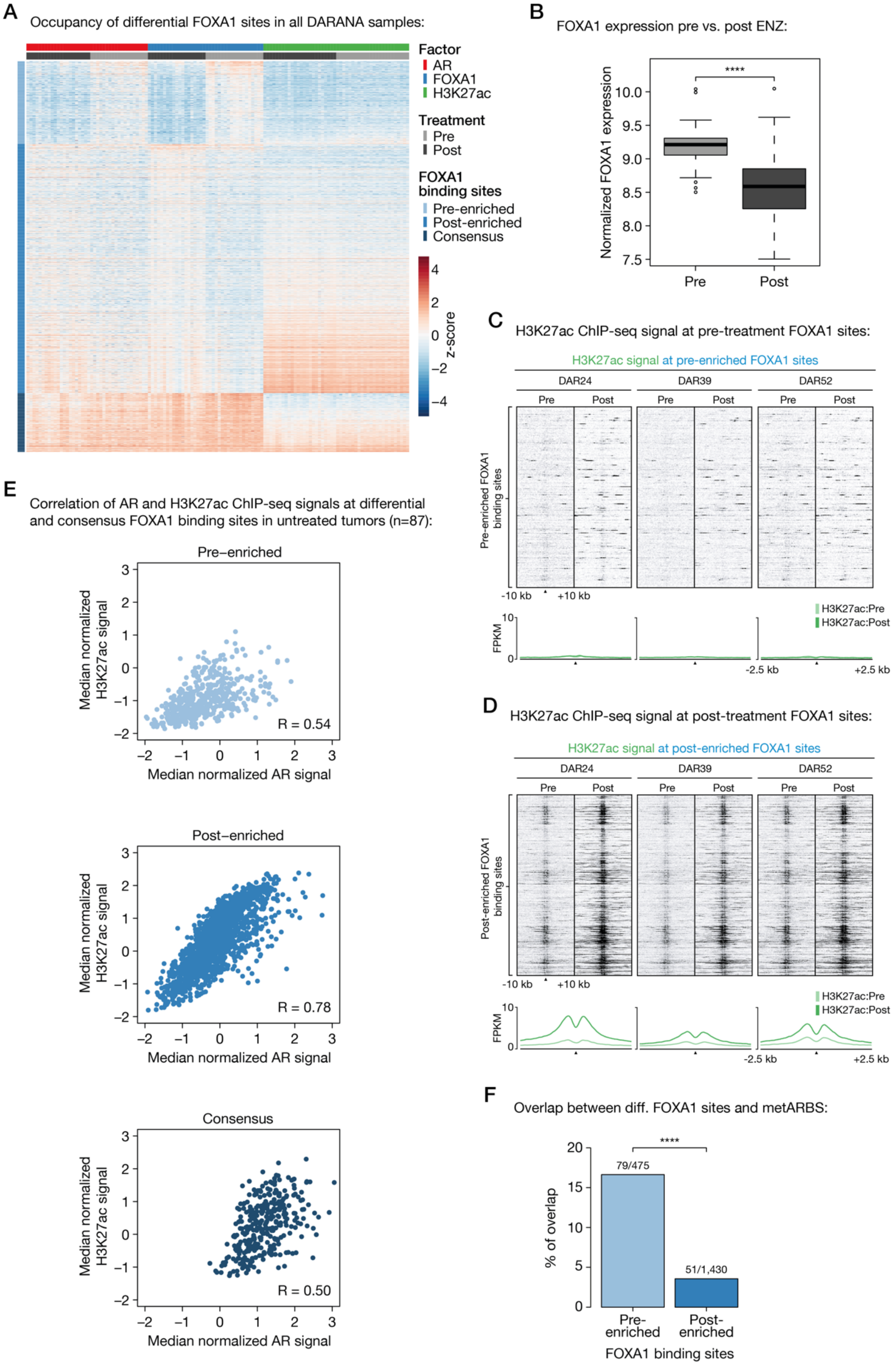


79

**Figure S6: Characterization of differential FOXA1 sites.**

(A) Coverage heatmap showing occupancy of differential (pre-/post-enriched) and consensus FOXA1 peaks in all generated pre- and post-treatment ChIP-seq samples. Heatmap color indicates region read counts (z-score) at pre-enriched, post-enriched and consensus FOXA1 sites (rows) in the pre- and post-treatment AR (red), FOXA1 (blue) and H3K27ac (green) ChIP-seq data streams (columns).

(B) Boxplot showing normalized FOXA1 gene expression before and after 3 months of neoadjuvant ENZ treatment. ****, $P < 0.0001$ (Mann-Whitney U-test).

(C-D) Representative tornado plots (top) and average density plots (bottom) visualizing H3K27ac ChIP-seq signal (in FPKM) at pre-enriched (C) and post-enriched (D) FOXA1 binding sites in 3 patients (DAR24, DAR39, DAR52) before and after ENZ treatment. Data are centered at FOXA1 peaks depicting a 10-kb (heatmaps) or 2.5-kb (density plots) window around the peak center.

(E) Scatter plots showing the correlation between AR and H3K27ac ChIP-seq signals at pre-enriched (top), post-enriched (middle) and consensus (bottom) FOXA1 sites in treatment-naïve primary prostate tumors (n=87). Pearson correlations for pre-enriched ($R = 0.54$), post-enriched ($R = 0.78$) and consensus ($R = 0.50$) FOXA1 sites are indicated.

(F) Bar chart indicating the overlap between pre-enriched (left) and post-enriched (right) FOXA1 binding sites, and previously identified (2) metastasis-specific AR binding sites (metARBS). ****, $P < 0.0001$ (Fisher's exact test).

#### Supplementary Figure S7

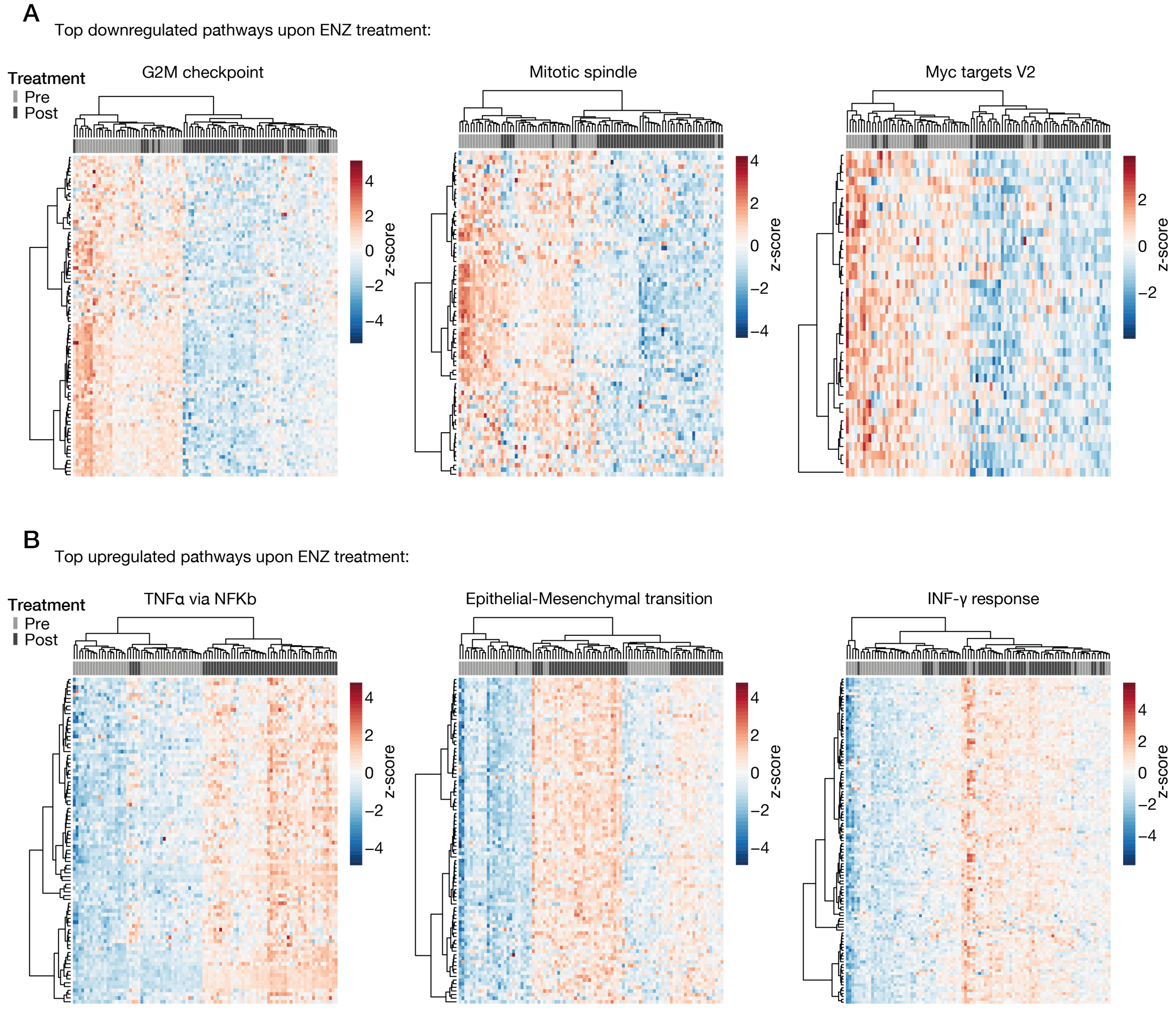


**Figure S7: Differential gene expression.**

Heatmaps depicting differential gene expression of top downregulated (A) and top upregulated (B) pathways upon ENZ treatment. Unsupervised hierarchical clustering of pre- and post-treatment RNA-seq samples is based on the expression of genes that define the respective hallmark gene sets. Color scale indicates gene expression (z-score).

#### Supplementary Figure S8

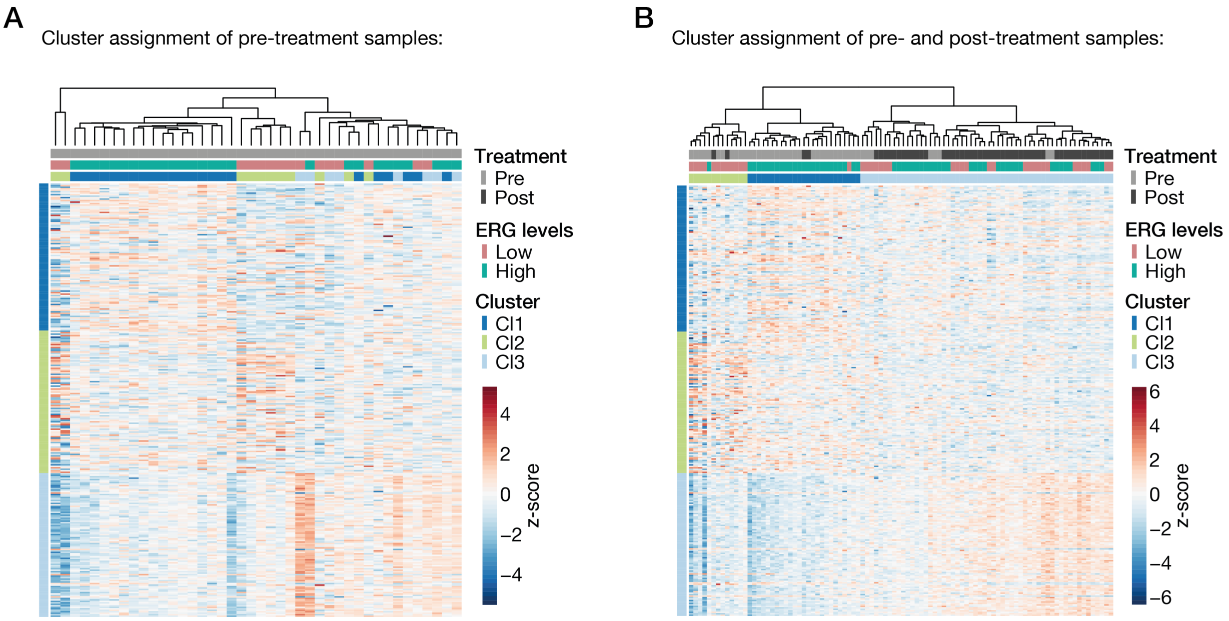


**Figure S8: Molecular subtyping.**

Unsupervised hierarchical clustering of pre-treatment (A) or pre- and post-treatment (B) RNA-seq samples using 285 genes differentially expressed across 3 previously reported PCa subtypes (3). For each sample, the assigned cluster affiliation as well as the ERG gene expression levels are indicated below the branching. The genes (rows) are ordered per cluster and the color scale of the heatmap indicates gene expression (z-score).

#### Supplementary Figure S9

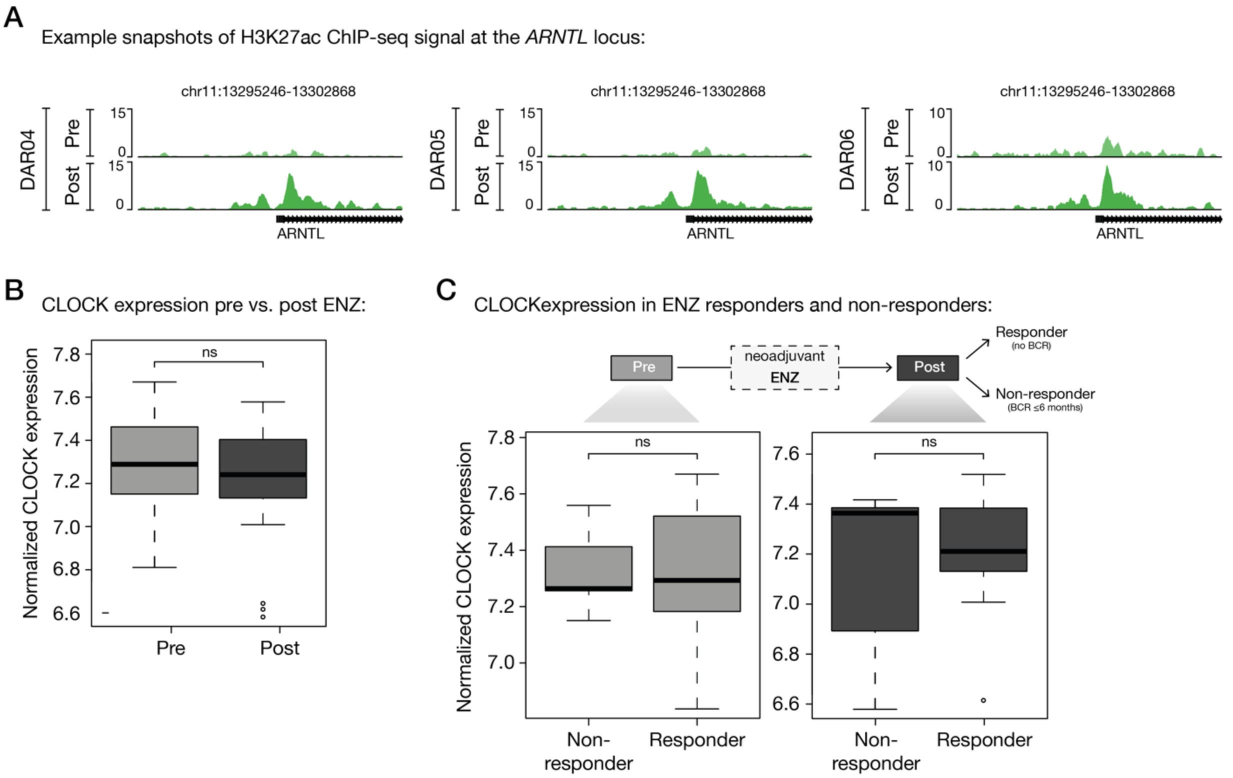


**Figure S9: Cistromic and transcriptomic profiling of circadian rhythm core components.**

(A) Representative example snapshots of H3K27ac ChIP-seq signal at the *ARNTL* gene locus before (top) and after (bottom) ENZ treatment. Shown are matched pre- and post-treatment ChIP-seq data of 3 patients (DAR04, DAR05, DAR06).

(B) Boxplot showing normalized CLOCK gene expression before and after 3 months of neoadjuvant ENZ treatment. ns, $P > 0.05$ (Mann-Whitney U-test).

(C) Boxplots depicting normalized CLOCK gene expression in ENZ non-responders (BCR ≤ 6 months; n=8) and responders (no BCR; n=29) in the pre- (left) and post-treatment (right) setting separately. ns, $P > 0.05$ (Mann-Whitney U-test).

#### Supplementary Figure S10

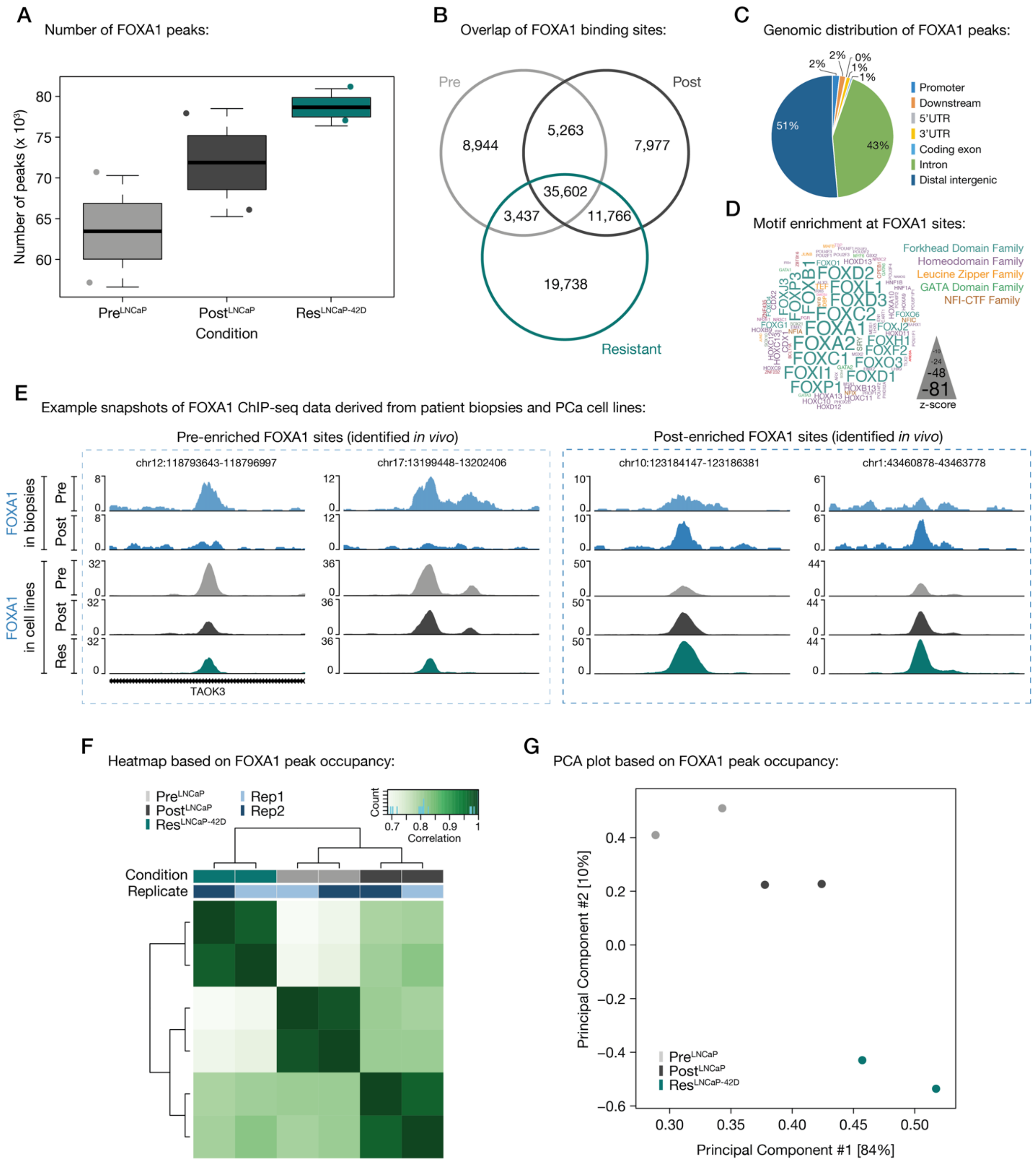


**Figure S10: Characterizing the FOXA1 cistrome in hormone-naïve and ENZ-resistant PCa cell lines.**

(A) Boxplot showing the number of FOXA1 peaks per ChIP-seq condition (Pre$^{LNCaP}$, Post$^{LNCaP}$, Res$^{LNCaP\text{-}42D}$). Peak calling was performed over matched cell line input control samples.

(B) Venn diagram indicating the overlap of FOXA1 binding sites in all tested cell line conditions ($\text{Pre}^{\text{LNCaP}}$, $\text{Post}^{\text{LNCaP}}$, $\text{Res}^{\text{LNCaP-42D}}$). For each condition, only peaks present in both replicates were included. The FOXA1 consensus cistrome across conditions (n=35,602 sites) was used for genomic distribution (C) and motif enrichment (D) analyses.

(C) Pie chart showing the genomic distribution of consensus FOXA1 binding sites.

(D) Word cloud shows motif enrichment at consensus FOXA1 sites. The font size represents the z-score and colors correspond to transcription factor families.

(E) Representative example snapshots of FOXA1 ChIP-seq signal at two pre-enriched (left) and two post-enriched (right) FOXA1 binding sites. Per genomic location, the pre- and post-treatment FOXA1 ChIP-seq signal from one patient (top), as well as the signal in all tested cell line models (bottom) is shown. For cell lines, the average of two biological replicates is represented. Y-axis indicates ChIP-seq signal in FPKM.

(F) Correlation heatmap based on FOXA1 peak occupancy. Clustering of the samples is based on all called peaks and represents Pearson correlations between individual ChIP-seq samples. The column color bars indicate the ChIP-seq condition ($\text{Pre}^{\text{LNCaP}}$, $\text{Post}^{\text{LNCaP}}$, $\text{Res}^{\text{LNCaP-42D}}$) and replicate information (Rep1, 2).

(G) Principal component analysis (PCA) plot based on FOXA1 peak occupancy. Each dot represents a ChIP-seq sample that is colored per condition (2 replicates per condition).

#### Supplementary Figure S11

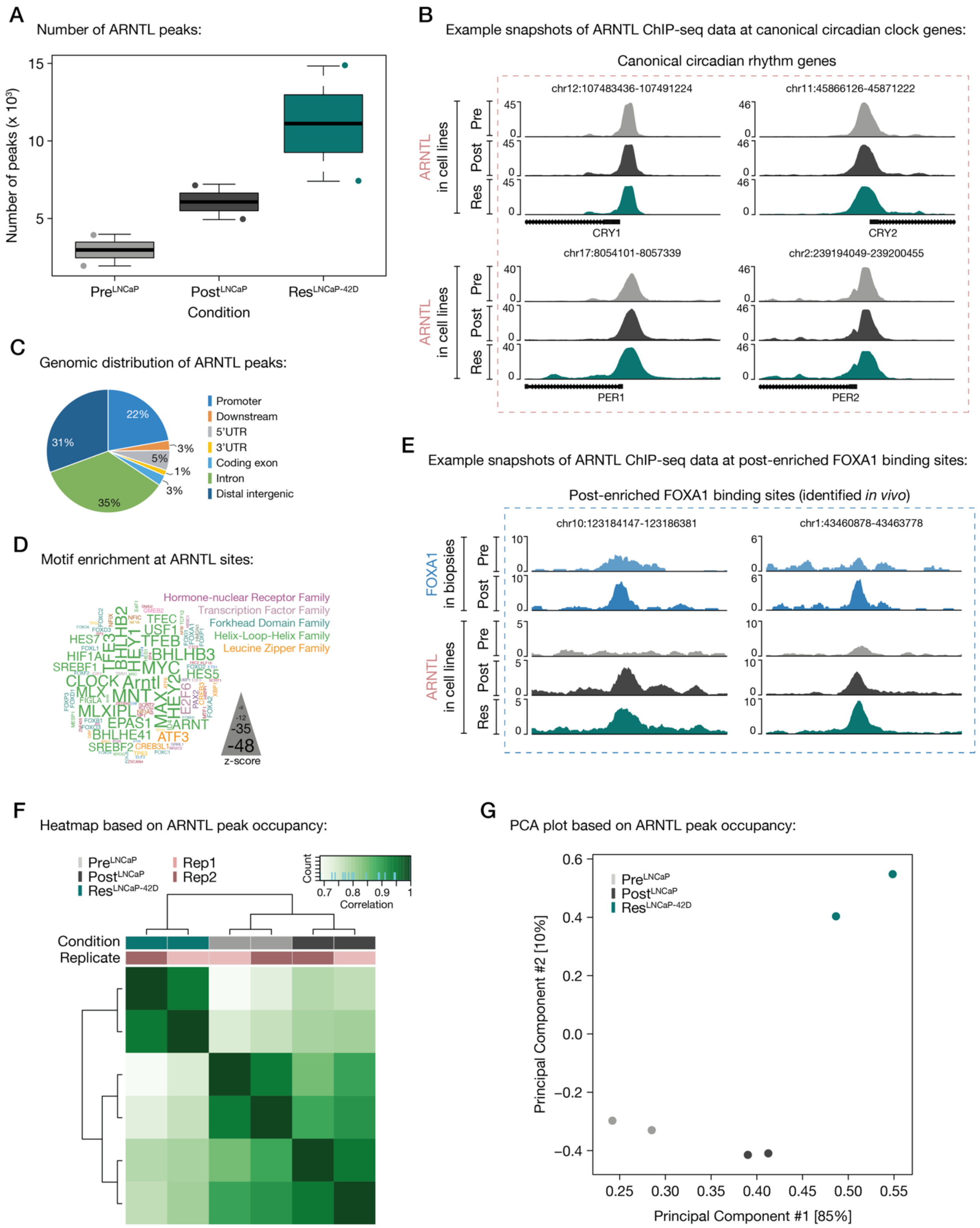


**Figure S11: Characterizing the ARNTL cistrome in hormone-naïve and ENZ-resistant PCa cell lines.**

(A) Boxplot showing the number of ARNTL peaks per ChIP-seq condition (Pre$^{\text{LNCaP}}$, Post$^{\text{LNCaP}}$, Res$^{\text{LNCaP-42D}}$). Peak calling was performed over matched cell line input control samples.

(B) Representative example snapshots of ARNTL ChIP-seq signal at canonical circadian rhythm genes. The average of two biological replicates is represented. Y-axis indicates ChIP-seq signal in FPKM.

(C) Pie chart showing the genomic distribution of ARNTL consensus sites (shared across all conditions; n=1,515 sites). The corresponding Venn diagram is shown in **Fig. 6E**.

(D) Word cloud shows motif enrichment at ARNTL consensus sites (n=1,515). The font size represents the z-score and colors correspond to transcription factor families. Since the human ARNTL motif is not part of the tested database, the homologous mouse motif (Arntl) was included.

(E) Representative example snapshots of ARNTL ChIP-seq signal at two post-enriched FOXA1 binding sites. Per genomic location, the pre- and post-treatment FOXA1 ChIP-seq signal from one patient, as well as the ARNTL signal in all tested cell line models (bottom) is shown. For cell lines, the average of two biological replicates is represented. Y-axis indicates ChIP-seq signal in FPKM.

(F) Correlation heatmap based on ARNTL peak occupancy. Clustering of the samples is based on all called peaks and represents Pearson correlations between individual ChIP-seq samples. The column color bars indicate the ChIP-seq condition (Pre$^{\text{LNCaP}}$, Post$^{\text{LNCaP}}$, Res$^{\text{LNCaP-42D}}$) and replicate information (Rep1, 2).

(G) Principal component analysis (PCA) plot based on ARNTL peak occupancy. Each dot represents a ChIP-seq sample that is colored per condition (2 replicates each).
